# Integrating enriched case data from national laboratory testing with population-based case-control analyses: a novel statistical likelihood-ratio methodology for PS4 applied to 325,345 breast cancer cases and 671,006 controls

**DOI:** 10.64898/2026.05.13.26353095

**Authors:** Sophie Allen, Charlie F. Rowlands, Alice Garrett, Fergus Couch, Marcy E. Richardson, Tina Pesaran, Joanna Pethick, Katrina Lavelle, Fiona McRonald, Sally Vernon, Beth Torr, Lucy Loong, Riyaad Aungraheeta, Miranda Durkie, George J. Burghel, Alison Callaway, Rachel Robinson, Joanne Field, Bethan Frugtniet, Sheila Palmer-Smith, Jonathan Grant, Judith Pagan, Trudi McDevitt, Katie Snape, Helen Hanson, Terri McVeigh, Chey Loveday, Michael Jones, Steven Hardy, Clare Turnbull, CanVIG-UK

**Author notes:** Corresponding author: Professor Clare Turnbull, Division of Genetics and Epidemiology, The Institute of Cancer Research, 32 Oakleaf Avenue, Sutton, SM2 5GP.

## Abstract

**Background:** For many evidence criteria within v3.0 of the ACMG/AMP guidelines, methodologies have been developed to empower their use outside the stipulated evidence strengths. However, no such methodology has been established for case-control data (PS4). With the release of large-scale unselected case-control datasets and expansion of nationally-collected laboratory datasets enriched for pathogenic variant carriers, there is potential to combine datasets across ascertainment contexts in a more quantitative manner using novel likelihood ratio tools.

**Methods:** Using our published PS4-LR-Calculator, we calculated a combined log likelihood ratio (PS4-LLR) across five datasets (three unselected, and two enriched), and estimated enrichment of pathogenic variants in clinically-ascertained laboratory data using truncating variant prevalence.

**Results:** Data were combined for 10,817 missense variants from 325,345 female breast cancer patients and 671,006 controls of Western European ancestry for five breast cancer susceptibility genes (*BRCA1, BRCA2, PALB2, ATM, CHEK2*). A combined LLR was produced for 4,690 missense variants; 927 variants received evidence towards pathogenicity (LLR≥ 1), and 3,242 received evidence towards benignity (LLR≤ -1).

**Conclusion:** This flexible, variant-level methodology combines nationally-collected ‘enriched’ datasets with unselected case-control cohorts, expanding the available information for case-control analysis, boosting power, enabling exploration of atypical penetrance and empowering variant classification.

## Background

The observation of a higher frequency of a variant of interest in cases of a given disease than in comparable controls is one of the most fundamental pieces of evidence by which it can be inferred that the variant is disease-associated and thus pathogenic. This evidence is especially valuable for missense variants for which, unlike protein truncating variants, the prior probability of pathogenicity is low. While individually rare, missense variants are in aggregate a common occurrence in diagnostic laboratories and frequently classified as variants of uncertain significance (VUS).

In the ACMG/AMP v3.0 framework, it was stipulated that case-control data of appropriate statistical significance (p<0.05) and appropriate effect size (OR≥5) could be awarded ‘strong’ evidence (PS4)(1). Unlike other evidence codes in the ACMG/AMP v3.0 framework, there was no subsequent specification by the ClinGen Sequence Variant Interpretation Group (SVI) of how more graded evidence might be awarded or how this frequentist threshold should be translated across to the Bayesian points-based framework they subsequently advocated in 2019 and which will be used across the v4.0 framework update due for release in 2027(2).

To address this, we devised a method using Gaussian adaptive quadrature (the PS4-LR-Calculator), by which, for a given set of observations of a variant in those with disease and those without disease, the likelihood that the variant is associated with disease is compared to the likelihood that the variant is non-associated, given a pre-specified target odds ratio for each hypothesis of association and non-association(3). This generates a likelihood ratio; the log (to base 2.08) of the likelihood ratio (LLR) can readily be applied as ‘evidence points’ within the ACMG/AMP Bayesian framework(4). LLRs can be readily summed across different case-control datasets, to maximise the evidence available for classification of a given variant. This type of approach has previously been illustrated in a recent analysis of variants in *BRCA1* and *BRCA2* in 96,691 unselected population-based cases of breast cancer and 302,116 controls from three case-control research datasets: BRIDGES, CARRIERS and UK Biobank(5–8).

However, there are a wealth of variant data from clinical diagnostic testing not captured when restricting to formal case-control research datasets. These clinical datasets firstly lack controls, and secondly include cases of diseases selected through application of eligibility criteria for testing. This unquantified level of “enrichment”, whilst potentially powerful in regard of variant interpretation, renders it challenging to then combine variant association metrics for these clinically-ascertained cases with those from population-based unselected breast cancer cases.

An additional emergent consideration is disease penetrance, which the PS4-LR-Calculator methodology is well-placed to address on account of its ability to flexibly accommodate any user-specified target odds ratio. The field of breast cancer susceptibility is ahead of most disease communities in having clinical consensus thresholds for defining genes of high penetrance (OR≥4, such as *BRCA1*, *BRCA2* and *PALB2*) and genes of moderate penetrance (OR≥2, such as *CHEK2* and *ATM*)(9). Accordingly, it is increasingly recognised that some variants in high penetrance genes are of reduced (moderate) penetrance and that some variants in moderate penetrance genes are of elevated (high) penetrance(10–13). This poses concomitant challenges in applying case-control data for variant classification.

Using the PS4-LR-Calculator, we have developed a case-control methodology for leveraging enriched nationally-collected breast cancer datasets in combination with unselected disease series. We present here combination of a clinical series of 187,642 enriched breast cases tested by Ambry, USA (analysed against 175,054 ancestry-matched controls from gnomAD v4.1.0(14)), a clinical series of 44,917 enriched breast cases tested by NHS laboratories in the UK (analysed against 297,141 ancestry-matched controls from UK Biobank) and 92,786 unselected breast cancer cases and 198,811 paired controls from BRIDGES, CARRIERS and UK Biobank. In total, we analysed a combined dataset of 325,345 white/European female breast cancer cases and 671,006 non-breast cancer/population controls for rare variants in *BRCA1, BRCA2, PALB2, CHEK2* and *ATM*, making this, to our knowledge, the largest case-control analysis of these genes to date. We have also investigated application of case-control data under reduced penetrance hypotheses for variants in genes of high penetrance, and high penetrance hypotheses for variants in genes of moderate penetrance.

## Methods

### Patients and data sources

Data was obtained for the *BRCA1*, *BRCA2, PALB2, ATM* and *CHEK2* genes from three population-based unselected breast cancer genetic testing studies (CARRIERS(7), UK Biobank(8), and 30 unselected studies from BRIDGES(6)) and from two enriched laboratory datasets (NDRS-UK, Ambry) (Table 1, Supplementary Methods)). We also included 175,054 non-UK Biobank non-Finnish European individuals from gnomAD v4.1.0 as population controls for use with the Ambry laboratory dataset(14) (Figure 1). For this analysis, ethnicity was restricted to white/European individuals in order to provide the largest possible available ethnicity-matched dataset of cases and controls from these constituent datasets, per the 2024 Association for Clinical Genomic Science (ACGS) UK recommendations for utilising case-control evidence(15).

**Table 1:** Summary of datasets included in analysis.

| Dataset | Genes | Ascertainment | Cases | Controls | Location | Cancer phenotype | Sex (cases) | Sex (controls) | Ancestry |
| --- | --- | --- | --- | --- | --- | --- | --- | --- | --- |
| **UK Biobank** | *ATM, BRCA1, BRCA2, CHEK2, PALB2* | Unselected | 18,477 | 419,373 | UK | Breast | Female | Female+ Male | White |
| **CARRIERS** | *ATM, BRCA1, BRCA2, CHEK2, PALB2* | Unselected | 32,247 | 32,544 | USA | Breast | Female | Female | Non-Hispanic White |
| **BRIDGES** | *ATM, BRCA1, BRCA2, CHEK2, PALB2* | Unselected | 42,062 | 44,035 | Europe | Breast | Female | Female | European |
| **Ambry** | *ATM, BRCA1, BRCA2, CHEK2, PALB2* | Clinical | 187,642 | NA | USA | Breast | Female | NA | White |
| **NDRS-UK** | *BRCA1, BRCA2* | Clinical | 44,917 | NA | UK | Breast | Female | NA | White |
| **gnomAD v4.1.0** | *ATM, BRCA1, BRCA2, CHEK2, PALB2* | Population | NA | 175,054 | Global | NA | NA | Female+ Male | Non-Finnish European |

**Figure 1:**
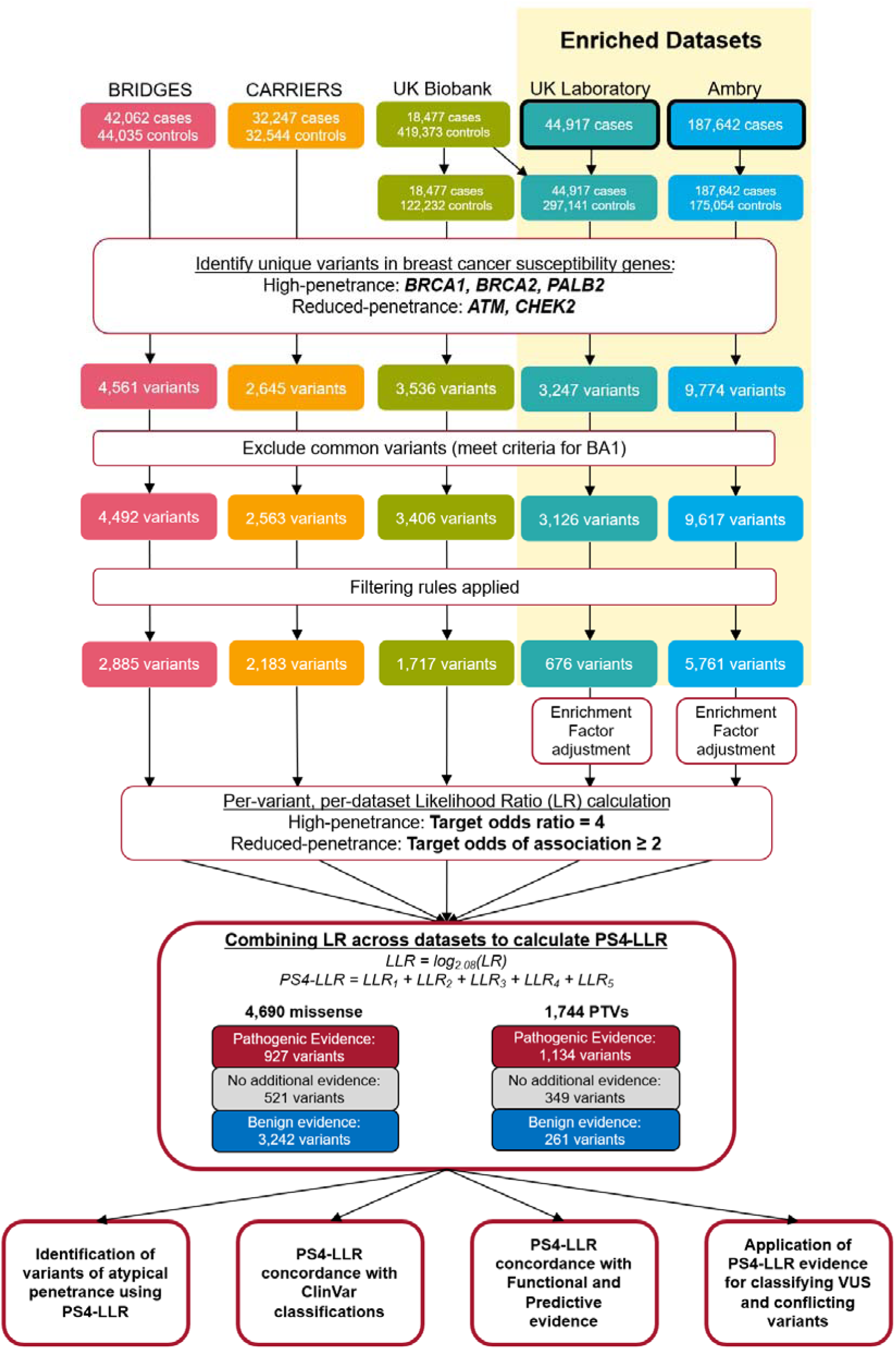
Overview of data collected and filtering steps employed to calculate PS4-LLR

Protein-truncating variants (PTVs) were defined as variants not in the final exon with one of the following variant effect consequences, annotated using Ensembl VEP(16) (v112): “stop_gained”, “splice_acceptor_variant”, “splice_donor_variant”, “frameshift_variant”. Missense variants were defined as variants with a VEP consequence of “missense_variant”. All variant nomenclature was verified using VariantValidator(17, 18).

For all datasets, variants which met the criteria for BA1 (allele frequency exceeding maximum tolerated allele frequency, MTAF) in gnomAD v4.1.0 were removed according to the MTAFs defined by the respective variant curation expert group (VCEP): *BRCA1*, 0.000868(13); *BRCA2*, 0.000906(13); *PALB2*, 0.00118(19); *ATM*, 0.00625(20); *CHEK2*, 0.00625(20).

### Case-control analysis

For each variant, the LR from each individual study was calculated using the previously published PS4-LR-Calc approach(3) (see Supplementary Methods for further details on this approach). Briefly, the calculator uses the observed frequency of a variant in both cases and controls to form two starting hypotheses: the hypothesis of association (that the underlying odds ratio indicates pathogenicity, the default threshold being greater than 5), hereafter ‘target odds of association’, and the hypothesis of non-association (that the underlying odds indicate benignity, the default threshold being lower than 1). The likelihood of each competing hypothesis being true is calculated by modelling the number of carriers in cases and controls as a binomial distribution, and the summed likelihood of each hypothesis compared to reach the final LR.

Initial review and analysis revealed the need for further rules to appropriately apply case-control evidence when using PS4-LR-Calc. These rules include removal of singleton observations, exclusion of benign evidence from laboratory datasets where benign variants were underreported, and calculation of an Enrichment Factor (EF) for enriched laboratory datasets (Supplementary Methods).

An observed odds ratio (OR), 95% confidence intervals, and Fisher’s exact p-value were also calculated for each variant in each dataset. An additional meta-analysis was carried out for the OR calculated for the three unselected datasets on a per-variant basis, using a fixed-effect inverse-variance weighted approach (Supplementary Methods).

All analyses were performed using R version 4.3.0 (2023-04-21 ucrt) and R Studio version 2025.05.1 Build 513. Kruskal-Wallis and Dunn tests were performed using the rstatix package for R. All plots were created using the ggplot2, ggpubr, and ggsankey packages for R.

### Combined LR calculation

Per-variant LR from each study were combined as follows:

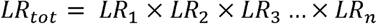

using data from n datasets for which an LR could be calculated. LR_tot_ were log transformed

(base 2.08) as per the Tavtigian *et al*. scaled point system to assess potential evidence strengths applicable for variant classification(2, 4):

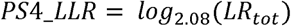

The default target odds of association used was 4 for high-penetrance genes (*BRCA1*, *BRCA2*, *PALB2*), and 2 for moderate penetrance genes (*ATM*, *CHEK2*), as per published clinical consensus recommendations for high increased risk and moderate increased risk for breast cancer relative to the general population risk(9, 13).

### Penetrance investigation

For high-penetrance genes (*BRCA1*, *BRCA2*, and *PALB2)*, four criteria were defined to identify potentially reduced penetrance variants when comparing evidence attained using the clinical consensus (OR≥4) and reduced penetrance (OR≥2) target odds of association:

- Evidence strength changes from benign to pathogenic
- Loses benign evidence (Benign Moderate or stronger to No Evidence)
- Gains pathogenic evidence (No evidence to Pathogenic Moderate or stronger)
- Substantially strengthens weakly pathogenic evidence (Pathogenic Moderate to Very Strong, or Pathogenic Supporting to Strong or Very Strong)

A change in evidence was defined as being a minimum change of two evidence strength categories (therefore a change of one category, e.g. supporting to moderate, would not meet any criterion)

For moderate-penetrance genes (*ATM* and *CHEK2)*, we highlighted putative high-penetrance variants by identifying variants which retained at least strong pathogenic evidence at OR≥4.

### Concordance of results with other data

Concordance of PS4-LLR was measured against 1 star or higher missense classifications from ClinVar (data release 01 June 2025)(21). For *BRCA1* and *BRCA2* variants, case-control evidence was compared against evidence applicable under functional (PS3/BS3) and predictive (PP3/BP4) criteria recommended by the ClinGen ENIGMA *BRCA1* and *BRCA2* Expert Panel Specifications to the ACMG/AMP Variant Interpretation Guidelines Version 1.2.0(13).

### Reinterpretation of missense VUS

Missense variants with classifications of VUS or conflicting interpretations in ClinVar were identified (data release 01 June 2025)(21). *BRCA1* and *BRCA2* variants were annotated with case-control evidence (in terms of PS4-LLRs) calculated using a target OR≥4.

Following the ClinGen ENIGMA *BRCA1* and *BRCA2* Guidelines, variants with results from high-throughput functional assays attained strong evidence for PS3 or BS3 (Supplementary Methods). Variants in potentially clinically important functional domains with data from BayesDel(22) and SpliceAI(23) exceeding the recommended thresholds attained supporting evidence for PP3 or BP4 (Supplementary Methods). Where variants met the above criteria, evidence was combined to attain an overall automated classification incorporating PS4-LLRs under the PS4 criterion. BA1, BS1, and PM2 were not used to avoid double-counting of population data already used in calculation of PS4-LLR.

## Results

### Dataset enrichment

Enrichment of pathogenic variants was estimated by comparing per-gene allele frequency of protein-truncating variants (PTV) in enriched cases against an average allele frequency of PTVs in unselected cases. The UK laboratory (NDRS-UK) dataset was found to be 4.84 times enriched for *BRCA1* PTVs and 3.92 times enriched for *BRCA2* PTVs. The Ambry dataset was also enriched to a lesser extent; all genes were between 1.81-2.63 times enriched in the Ambry dataset (Table 2, Supplementary Table 1). These enrichment factors were used to upscale the target odds of association in calculation of PS4-LLRs in respective enriched datasets (see Supplementary Table 14).

**Table 2:**
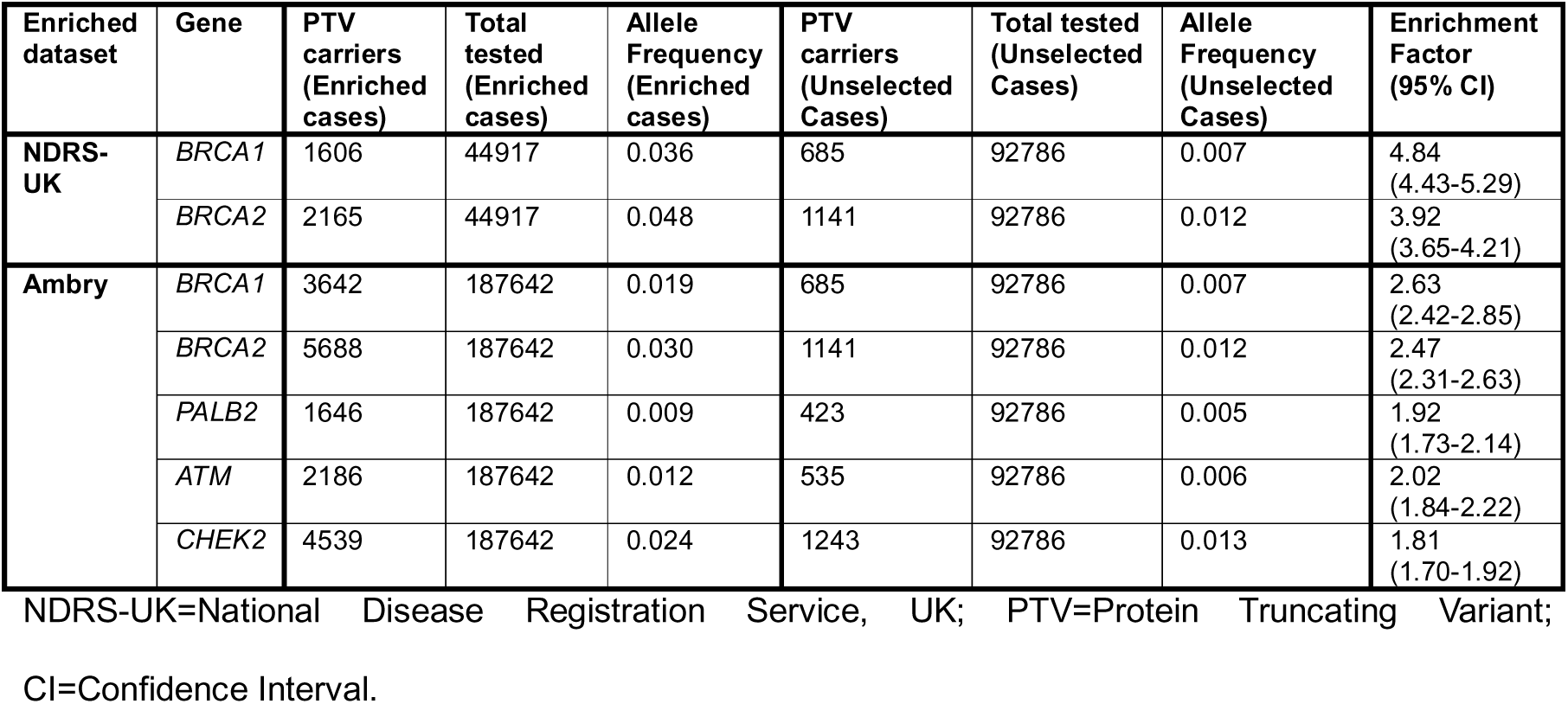
Per-gene enrichment factors calculated by comparison of truncating variant prevalence.

### PS4-LLR results from combined dataset

In total, 13,966 rare PTV and missense variants in five genes were identified across the five datasets (in 325,345 female breast cancer cases and 671,006 controls) (Supplementary Table 2).

Of 3,149 rare PTVs, 45% (1,405/3,149) were excluded from analysis due to violating at least one variant inclusion rule (Supplementary Methods), of which 96% (1,350/1,405) were due to being singleton observations (Supplementary Table 3). Of the remaining 1,744 variants, 65% (1,134/1,744) attained PS4-LLR evidence towards pathogenicity (314, 181, 274, and 365 at supporting, moderate, strong, or very strong PS4-LLR respectively), and 15% (261/1,744) attained evidence towards benignity (40, 89, 79, and 53 at supporting, moderate, strong, or very strong respectively).

Of 10,817 rare missense variants, 57% (6,127/10,817) were excluded from analysis, 91% (5,570/6,127) due to being singleton observations (Supplementary Table 4). Of the remaining 4,690 missense variants, 20% (927/4,690) attained PS4-LLR evidence towards pathogenicity (LLR≥1), 69% (3,242/4,690) attained PS4-LLR evidence towards benignity (LLR≤-1) and 11% (521/4,690) attained no evidence towards pathogenicity/benignity.

Considering the ACMG/AMP v3.0 evidence categories of supporting, moderate, strong, very strong, 84% (485/579) of *BRCA1* missense variants with benign evidence (LLR≤-1) attained at least strong evidence (LLR≤-4), while 39% (49/127) of *BRCA1* variants with pathogenic evidence (LLR≥1) attained at least strong evidence (LLR≥4). Similarly, for *BRCA2*, 82% (957/1,164) and 38% (80/211) variants reached at least strong evidence towards benignity and pathogenicity respectively. By comparison, far fewer *PALB2* variants attained strong/very strong pathogenic evidence (14%; 12/85, Table 3).

**Table 3:** Overview of evidence attained for 4,690 rare missense variants included in the combined dataset.

| | TOTAL VARIANTS | Any pathogenic evidence (% total) | No evidence (% total) | Any benign evidence (% total) | PS4-LLR Evidence towards pathogenicity ($LLR \geq 1$) | | | | PS4-LLR Evidence towards benignity ($LLR \leq -1$) | | | |
| --- | --- | --- | --- | --- | --- | --- | --- | --- | --- | --- | --- | --- |
|  |  |  |  |  | VSTR | STR | MOD | SUP | SUP | MOD | STR | VSTR |
| *BRCA1* | 808 | 127 (16) | 102 (13) | 579 (71) | 25 | 24 | 35 | 43 | 22 | 72 | 159 | 326 |
| *BRCA2* | 1570 | 211 (13) | 195 (13) | 1164 (74) | 26 | 54 | 59 | 72 | 52 | 155 | 362 | 595 |
| *PALB2* | 561 | 85 (15) | 47 (8) | 429 (77) | 4 | 8 | 35 | 38 | 16 | 88 | 112 | 213 |
| *ATM* | 1375 | 364 (26) | 135 (10) | 876 (64) | 24 | 61 | 125 | 154 | 87 | 254 | 290 | 245 |
| *CHEK2* | 376 | 140 (37) | 42 (11) | 194 (52) | 25 | 27 | 28 | 60 | 12 | 63 | 66 | 53 |
| **TOTAL** | **4690** | **927 (20)** | **521 (11)** | **3242 (69)** | **104** | **174** | **282** | **367** | **189** | **632** | **989** | **1432** |

Target odds of association were set at $OR \geq 4$ for *BRCA1*, *BRCA2*, and *PALB2*, and $OR \geq 2$ for *ATM* and *CHEK2*. Evidence strengths are provided as follows for each gene: VSTR=Very Strong, STR=Strong, MOD=Moderate, SUP=Supporting

For moderate penetrance genes, assessed against a target OR of 2, 23% (85/364) and 61% (535/876) of *ATM* variants with evidence towards pathogenicity and benignity respectively attained at least strong evidence. For *CHEK2*, the proportion of variants which attained evidence towards pathogenicity was higher than any other gene at 37% (140/376); of these, 37% (52/140) attained at least strong evidence.

Examining PS4-LLR as continuous scores across each gene (not divided into ACMG/AMP v.3.0 categories), evidence ranged from +153 to -540, with *PALB2* having the narrowest evidence range (+16 to -176). Extreme PS4-LLRs towards pathogenicity were observed for relatively frequent well-established pathogenic variants; notably, *BRCA1*: c.181T>G (p.C61G) (+152), *BRCA2*: c.7988A>T (p.E2663V) (+139), c.8167G>C (p.D2723H) (+125), *ATM*: c.7271T>G (p.V2424G) (+65) (see Discussion).

Median PS4-LLR for each gene was negative (i.e. LLR<0), demonstrating that overall, as might be anticipated, more evidence was attained by variants towards benignity than pathogenicity (Figure 2). Median PS4-LLR for *BRCA1*, *BRCA2,* and *PALB2* were -5.11, - 5.54, and -5.34. Comparing distributions of PS4-LLR for each gene, average PS4-LLR for *ATM* and *CHEK2* differed significantly from *BRCA1*, *BRCA2*, and *PALB2* (Figure 2).

**Figure 2:**
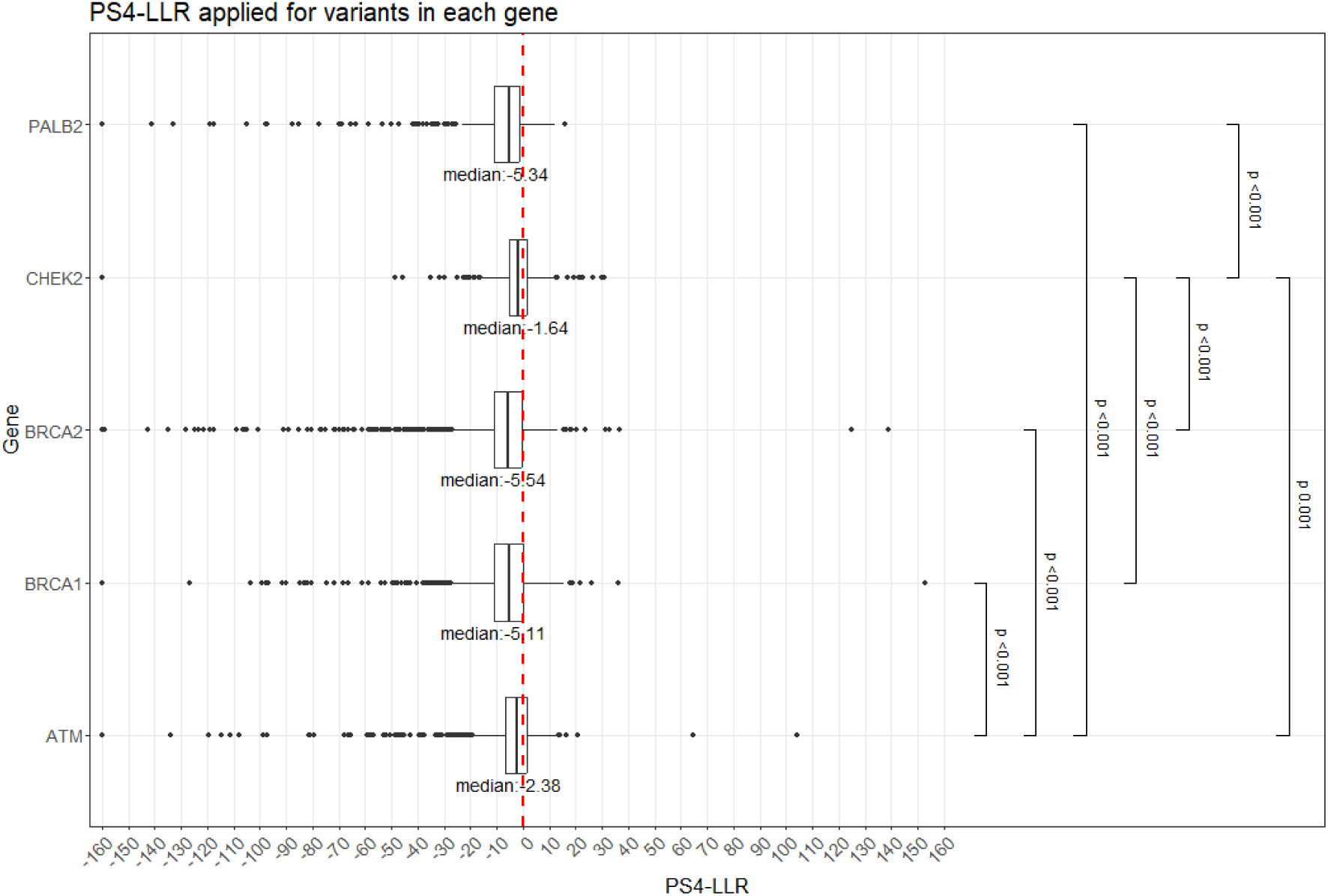
Boxplot of PS4-LLR evidence assigned for 4,690 rare missense variants included in the combined dataset. Each dot represents a single variant. Red dashed line represents 0 evidence points attained. For 28 variants with negative PS4-LLR more extreme than -160, LLR was set to a maximum limit of -160. Statistically significant differences between average LLR for each gene are displayed (Kruskal-Wallis rank sum: p-value=3.73×10^-57^, Bonferroni-adjusted p-value threshold for significance=0.005)

### Reduced penetrance variants in high penetrance genes

In addition to calculating PS4-LLR at the clinical consensus target odds ratios, we examined the change in applicable evidence strengths at different target odds ratios to reflect atypical penetrance scenarios. For genes with higher breast cancer penetrance (*BRCA1*, *BRCA2* and *PALB2)*, we compared PS4-LLRs at target odds of association of OR≥4 to OR≥2 for the context of reduced penetrance.

Of the 2,939 rare missense variants in these three genes, the shift in PS4-LLR when reducing the target odds ratio from 4 to 2 indicated possible reduced penetrance for 427 variants (14.2% (115/808) in *BRCA1*, 13.9% (219/1570) in *BRCA2,* and 16.6% (93/561) in *PALB2* (Supplementary Fig. 3, Supplementary Table 5, see Methods), of which 56% (240/427) notably already had a conflicting classification in ClinVar.

### High penetrance variants in (otherwise) moderate penetrance genes

For genes with moderate breast cancer penetrance (*ATM* and *CHEK2*), we examined the impact of increasing the target odds of association from OR≥2 to OR≥4 for variants which had attained at least very strong pathogenic evidence at a target odds of association of 2 (the clinical consensus odds ratio for these genes as suggested by the ClinGen HBOPC VCEP for *ATM*(*20*)) (Supplementary Table 6).

37 rare missense variants in *ATM* and *CHEK2* maintained at least strong evidence at this increased target odds ratio of OR≥4: 1.5% (21/1,375) in *ATM* and 4.3% (16/376) in *CHEK2*. Notably included in this list was *ATM* c.7271T>G (p.V2424G), a well-established high-penetrance dominant-negative variant.

### Concordance with ClinVar classifications

Of the total of 26,629 missense variants listed in ClinVar for *BRCA1, BRCA2, PALB2, CHEK2* and *ATM* and reported with at least 1 star, 1,214 were reported as Pathogenic, Likely Pathogenic, Likely Benign, or Benign, and 25,415 variants were reported as VUS or with conflicting classifications. 17% (4,555/26,629) of missense variants in ClinVar have corresponding data in our final combined dataset; 27% (323/1,214) of those with at least 1 star, and 17% (4,231/25,415) of VUS or conflicting variants.

For the 323 classified missense variants with at least 1 star, 71% (85/120) of ClinVar P/LP classified variants attained PS4-LLR towards pathogenicity (LLR≥1), with 9% (11/120) attaining no points and 20% (24/120) attaining PS4-LLR points towards benignity with 2, 8, 9, and 5 variants in the supporting, moderate, strong and very strong range respectively. These 24 discordant variants include known reduced penetrance variant *BRCA1* c.5096G>A p.R1699Q, and 4 variants in this group were additionally flagged as potentially of reduced penetrance when using a target odds of association of OR≥2 (*BRCA2* c.7879A>T p.I2627F, *BRCA2* c.8177A>G p.Y2726C, *BRCA2* c.9302T>G p.L3101R, *PALB2* c.3350G>A p.R1117K).

88% (178/203) of ClinVar B/LB classified variants attained PS4-LLR towards benignity (LLR≤-1), with 6% (12/203) attaining no points and 6% (13/203) attaining PS4-LLR towards pathogenicity with 5, 4, 3, and 1 variants respectively in the supporting, moderate, strong, very strong range (Figure 3, Supplementary Table 7).

**Figure 3:**
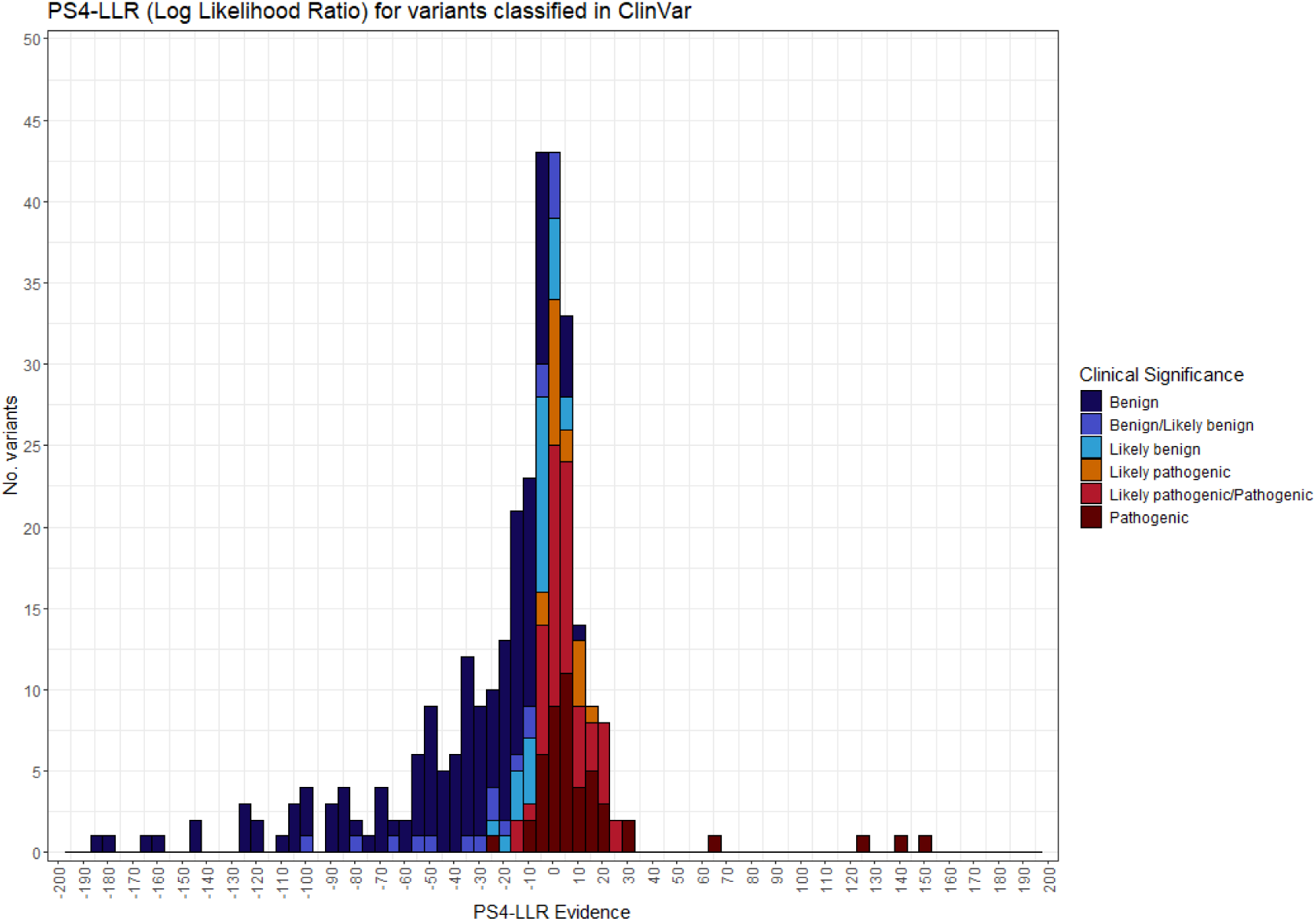
323 missense variants across five genes with PS4-LLR between 200 and -200. Missense variants included were classified in ClinVar as either pathogenic, likely pathogenic, likely benign, or benign with at least one star. 13 variants with a PS4-LLR≤-200 not depicted; all 13 of these variants were classified as Benign with 3* expert panel classifications. PS4-LLR was calculated using OR≥4 for *BRCA1*, *BRCA2*, and *PALB2*, and OR≥2 for *ATM* and *CHEK2*.

To minimise impact of single submitters to ClinVar, which may be historic and/or erroneous, we repeated this analysis restricting inclusion of ClinVar records to those with at least 2 stars (multiple lab submissions and/or expert panel classifications) (Supplementary Fig. 2). 146 *BRCA1* and 126 *BRCA2* variants met this criteria. Of these 272 variants, 77% (60/78) of ClinVar 2 star (and above) pathogenic variants attained concordant PS4-LLR evidence towards pathogenicity, with 15% (12/78) attaining PS4-LLR evidence towards benignity with 1, 3, 6, and 2 in the supporting, moderate, strong and very strong range respectively. 89% (173/194) of benign variants with PS4-LLR attained concordant evidence towards benignity, with 5% (10/194) attaining PS4-EPs towards pathogenicity with 4, 2, 3, and 1 respectively in the supporting, moderate, strong, very strong range.

### Concordance with functional data and in silico predictions

Of 2,030 *BRCA1/BRCA2* variants with a VUS or conflicting classification in ClinVar, 253 had at least supporting evidence from both functional data and case-control data (156 in *BRCA1* and 97 in *BRCA2*). For 87% (220/253), the functional evidence was concordant with the direction of evidence from case-control data (87% (135/156) in *BRCA1* and 88% (85/97) in *BRCA2*). There were 24 discordant variants with functional evidence indicating neutrality, with 7, 5, 10, and 2 respectively attaining PS4-LLR evidence towards pathogenicity in the supporting, moderate, strong and very strong range. There were 9 discordant variants with functional evidence of deleteriousness, with 0, 3, 3, and 3 respectively attaining PS4-LLR evidence towards benignity in the supporting, moderate, strong, very strong range. Of these 9 variants, 3 variants were additionally flagged as potentially of reduced penetrance (*BRCA1* c.4988T>A p.G1763V, *BRCA2* c.7871A>G p.Y2624C, *BRCA2* c.7786G>A p.G2596R)

For the 336 *BRCA1/BRCA2* variants with at least supporting evidence from both in silico predictive tools (BayesDel, SpliceAI) and case-control data, (92 in *BRCA1* and 244 in *BRCA2*), 74% (249/336) had predictive evidence that was concordant with the direction of evidence from case-control data (69% (64/92) in *BRCA1* and 76% (185/244) in *BRCA2*). There were 37 discordant variants with predictive evidence indicating neutrality, of which 12, 13, 8, and 4 respectively attained PS4-LLR towards pathogenicity in the supporting, moderate, strong, and very strong ranges. There were 51 discordant variants with predictive evidence of deleteriousness, of which 2, 8, 19, and 22 respectively attained PS4-LLR towards benignity in the supporting, moderate, strong, and very strong ranges. 18% (9/51) of these discordant variants were additionally flagged as potentially of reduced penetrance when using a target odds of association of OR≥2 to calculate PS4-LLR (Supplementary Table 8, Supplementary Table 5)

### Application of PS4-LLR evidence for classifying VUS and conflicting variants

In total, 89% (3,767/4,231) of VUS and conflicting variants present in our five datasets have PS4-LLR applied towards either pathogenicity or benignity, and of these, 54% (2,030/3,767) have this evidence applied at strong or higher (Supplementary Table 7). These variants might benefit from quantified case-control evidence to enable classification out of their VUS status.

For the 2,030 VUS or conflicting variants with case-control evidence in *BRCA1* and *BRCA2*, 531 also had functional and/or predictive evidence. To meet the ACMG/AMP v3.0 requirement that classifications are reached using at least two non-contradicting criteria(1), variants were only classified if they had case-control evidence plus either concordant functional data or predictive data.

19 variants currently ClinVar-assigned as VUS (12 in *BRCA1*, 7 in *BRCA2*) attained at least Likely Pathogenic using these evidence items, and 382 (147 in *BRCA1*, 235 in *BRCA2*) variants attained at least Likely Benign. This totalled 76% (401/531) of variants which attained sufficient evidence for classification using automated evidence application including quantified case-control data (Supplementary Table 8).

## Discussion

We present here the largest case-control analysis to date involving data from 325,345 breast cancer cases and 671,006 controls, examining PS4-LLR in five genes and generating evidence assisting classification for 6,434 rare variants (4,690 rare missense variants). We focus here on missense variants, as this variant type stands to benefit most towards definitive classification from a more quantitative approach to case-control data. PS4-LLRs are also presented for protein truncating variants (PTVs) (Supplementary Table 2); the average per-gene evidence towards pathogenicity is greater for protein truncating variants compared to missense (Supplementary Fig. 1) but these variant types tend to achieve classification more readily by virtue of high weight placed on mutational type.

### Patterns of PS4-LLR scores across and between genes

57% (6,127/10,817) of missense variants could not be included for PS4-LLR analysis, nearly all due to being singleton observations. Of the 4,690 recurrently-observed analysable variants, overall 20% (927/4,690) of variants received PS4-LLR evidence towards pathogenicity (LLR≥1) and 69% (3,242/4,690) received PS4-LLR evidence towards benignity (LLR≤-1).

Observed differences between the genes in variant numbers will in part reflect inherent characteristics regarding gene length and constraint, as well as the greater volume of testing data for *BRCA1/BRCA2*. A variant will more readily attain evidence towards benignity in the context of application of a higher target OR, meaning that the distribution of benignity scores is accordingly skewed for *BRCA1, BRCA2* and *PALB2* compared to *ATM* and *CHEK2*. Furthermore, even amongst our high-penetrance genes, although measured against a standardised working clinical definition of high penetrance of OR≥4, the magnitude of their underlying association with breast cancer differs quite widely between genes, estimated in a recent large meta-analysis as being OR≥8.73 for *BRCA1*, OR≥5.68 for *BRCA2* and OR≥4.30 for *PALB2*(24). Thus, a *BRCA1* missense variant of ‘true underlying OR’ typical for a *BRCA1* PV will attain stronger PS4-LLR towards pathogenicity than a *BRCA2* missense variant of ‘true underlying OR’ typical for *BRCA2* PV when both are measured against a target OR≥4. Consistent with this, we observed for *BRCA1* a higher proportion of variants attaining points towards pathogenicity and a higher proportion attaining strong/very strong than *BRCA2*. Also consistent with this, we observed a lower proportion of *PALB2* variants attaining strong/very strong PS4-LLR towards pathogenicity (although it has also been posited from other analyses that very few *PALB2* missense variants are pathogenic)(19). While we present PS4-LLR calculated using OR≥4 to align with current standard clinical classification practice for these genes, we also present as a comparison PS4-LLR calculated using bespoke target odds for each gene (as estimated from the recent meta-analysis; Supplementary Table 12).

Variant frequency also influences scores owing to increased power. The *BRCA2* variant c.1786G>C (p.D596H) illustrates attainment of a sizeable benignity score of -540 (Supplementary Table 2) through combination of a comparatively high variant frequency (observed 593 times in controls in UKB) and the higher target odds ratio used for *BRCA1* compared to *ATM/CHEK2*. Variant frequency (power) likewise boosted pathogenicity scores for some highly recurrent variants, such as *BRCA1*: c.181T>G (p.C61G) (+152), *BRCA2*: c.7988A>T (p.E2663V) (+139) and c.8167G>C (p.D2723H) (+125).

For the moderate penetrance genes *CHEK2* and *ATM,* the underlying odds ratio is more similar to the operational target odds ratio of 2, being estimated in recent meta-analysis as being 2.16 for *ATM* and 2.40 for *CHEK2*(*24*). While a lower proportion of *ATM* missense variants attained the highest category of PS4-LLR score (very strong), and the median score among all scored variants in *ATM* was lower than for *CHEK2* (−2.38 versus -1.64), the range of scores for *ATM* was much broader than for *CHEK2* (Figure 2). This likely reflects inclusion of some more recurrent high-scoring variants such as *ATM*: c.6919C>T (p.L2307F) (+104) and c.7271T>G (p.V2424G) (+65). Interestingly, whilst *ATM* c.7271T>G (p.V2424G) is a known high-penetrance pathogenic variant, *ATM* c.6919C>T (p.L2307F) has previously been classified as being benign on the basis of its high frequency in the Ashkenazi Jewish (AJ) population in gnomAD v2.1.1; this sizeable case-control signal would suggest a possible founder effect in the AJ population rather than neutrality of the variant, and indeed this variant has previously been associated with other cancers in AJ populations(25–27).

### Inclusion of enriched clinical datasets: adjustments and utility

We illustrate for the first time combination of enriched datasets with unselected datasets to generate an integrated likelihood ratio scoring. Notably the NDRS-UK laboratory data were more enriched for PTVs in *BRCA1* and *BRCA2* compared to the Ambry dataset, reflecting the stricter criteria for testing eligibility. It was also interesting to note the different enrichment factors by gene, which are a predictable reflection of how subtype-specific breast cancer association varies by gene. This, along with differences in the case make-up of datasets due to eligibility for diagnostic testing (for example, disproportionate inclusion of triple-negative breast cancers), are consistent with the higher enrichment factor for *BRCA1*.

Inclusion of the Ambry-US and NDRS-UK diagnostic laboratory datasets greatly increased power, adding an additional 232,559 enriched breast cancer cases to the 92,786 available from the three unselected series, and resulting in potentially applicable evidence for 4,563 missense variants, compared with 926 available missense variants if using case-control data from only the three unselected datasets using a meta-analysis (Table 3, Supplementary Table 9).

### Using PS4-LLRs to identify variants of atypical penetrance

Our existing clinical approaches to variant classification are based on a simplistic assumption that the variant will be either “fully penetrant” (in quasi-Mendelian fashion) or have no association with disease. We are as a community recognising an urgent requirement for new approaches to identification and classification of variants of reduced penetrance, not least as these will require different clinical management.

Analysing all variants in high penetrance genes *BRCA1*, *BRCA2* and *PALB2* against both a target odds ratio of 4 (high penetrance) and 2 (moderate penetrance), we were able to flag 427 variants as being of putatively-reduced penetrance, 240 of which also had a conflicting classification in ClinVar. These included *BRCA2* c.9302T>G (p.L3101R), a variant previously asserted as being of reduced penetrance based on a cancer family history algorithm(28) (Supplementary Table 5, Supplementary Table 12). However, this analysis did not identify all known reduced penetrance missense variants; interestingly, *BRCA1* c.5096G>A (p.R1699Q)(29), proposed and clinically-managed as reduced penetrance on the basis of segregation analyses and reported as Pathogenic based on expert panel submission to ClinVar, retained strong scoring for benignity against target ORs of either 4 or 2. This was driven by gnomAD counts in the Ambry dataset, and supported by the complete absence of the variant in breast cancer cases in CARRIERS and UK Biobank. On previous case-control analysis of this variant in unselected datasets, an odds ratio towards benignity was likewise reported, indicating this to be a product of the constituent datasets rather than methodology(5).

We also identified 37 variants in moderate penetrance genes that are putatively of high penetrance: these included the one well-established exemplar variant of this class *ATM* c.7271T>G (p.V2424G), which has long been recognised to be of atypically high penetrance and is managed clinically as per a *BRCA1/BRCA2* variant(30).

### Concordance of PS4-LLR scores with other data sources

Overall concordance against previously classified missense variants in ClinVar for all genes was relatively high, with 71% (85/120) P/LP ClinVar classified variants attaining PS4-LLR≥1 (evidence towards pathogenicity), and 88% (179/204) of B/LB ClinVar classified variants attaining PS4-LLR≤-1 (evidence towards benignity). Concordance was modestly higher when restricting to 2* classifications (77% and 89% for PS4-LLR towards pathogenicity and benignity respectively). Notably, very few *ATM*, *CHEK2* or *PALB2* missense variants attained 1* or higher ClinVar classifications, meaning assessment of concordance was mostly based on *BRCA1* and *BRCA2* ClinVar classifications.

Where discordant with ClinVar, typically variants only attained supporting or moderate PS4-LLR evidence in the discordant direction. Some discordant data is expected between ClinVar and PS4-LLR evidence as PS4-LLR represents a single evidence component, while ClinVar classifications incorporate the full spectrum of applicable evidence, and additionally PS4-LLR is impacted by low and stochastic frequency in case datasets. However, one plausible explanation for a proportion of the discordancies will be variants being of reduced penetrance, meaning these variants attain PS4-LLR evidence towards benignity when assessed against a high penetrance target OR (Supplementary Table 13). Notably for PTVs, 66 *BRCA1, BRCA2*, and *PALB2* variants with a pathogenic/likely pathogenic ClinVar classification attained at least strong benign evidence at OR≥4, including suspected reduced penetrance variant *BRCA2* c.8331+2T>C (Supplementary Table 13). It should also be noted that these data do not account for variability in genotype-phenotype associations (for example a *BRCA1* variant conferring hypothetical “ovarian-predominant, breast-minimal” risk). As such, the PS4-LLR variant evidence presented is a reflection only on breast cancer risk.

Although majority concordant with functional and in silico predictions, discordancies with these data may again highlight reduced (or alternative) penetrance. Indeed, several variants flagged in this analysis were found to have discordant evidence when compared against functional and/or in silico predictions. Additionally, functional and in silico data types are not ‘gold-standards’ for classification status. Functional classifications, as per the Brnich guidance, are assigned dichotomously, meaning that some variants assigned as functionally deleterious may exbibit weaker assay scores and lie very near to the assigned threshold for deleteriousness. In silico tools are widely recognised to over-predict deleteriousness, which was consistent with the pattern we observed in our discordancies.(5, 31, 32)

### Limitations

To avoid stratification due to population structure we restricted our analyses to the white-only subsets of these datasets. With growing availability of additional datasets containing sizeable numbers of non-white participants such as AllOfUs, application of this methodology may generate PS4-LLR scores for additional variants for which allele frequencies are higher in non-white ancestry groups.

A feature of the PS4-LLR methodology is that when variants have well-powered association signals lying between OR≤1 and the target OR, the likelihood areas for ratio comparison can be excessively small precluding robust analysis: for these reasons a handful of relatively frequent alleles well-established as having lower-penetrance breast cancer association will not be included in this analysis, such as *CHEK2* c.470T>C (p.I157T)(33) (Supplementary Table 14).

For BRIDGES and CARRIERS, the control series were female only. In order to optimise the case-control balance for matching of controls to the clinical series, it was necessary to retain male samples in the control series of UK Biobank and gnomAD v4.1.0. In principle, this might potentially have resulted in a very small degree of attenuation of the case-control signal, which would serve only to dampen very slightly the LLR towards pathogenicity.

For the NDRS-UK diagnostic laboratory data, laboratories only submitted rare variants classified internally as VUS, pathogenic or likely pathogenic. Because variants were not submitted by the laboratories if presumptively benign, the case counts for some variants may be under-represented. Thus, we excluded evidence attained towards benignity from this dataset. Additionally, we acknowledge that heterogeneity in variant calling methods between datasets means absence of a variant in a dataset may reflect lower coverage of that region in that dataset. For both of these reasons which reduce the number of informative datasets contributing to PS4-LLR, we recognise that the scoring towards pathogenicity may have been conservative, underscoring some of the caveats of using multi-centre real-world healthcare data.

We only examined data for five breast cancer susceptibility genes, and only considered the two ‘clinically-actionable’ thresholds for effect sizes. It will be of value to explore application of the likelihood ratio methodology to a broader range of gene-disease paradigms, for example exploring rare disease scenarios in which a much higher effect size would be anticipated (ie OR≥100 or OR≥1000). Quantitative harnessing of case-control data is important not only for generation of direct evidence for variant interpretation, but can afford accurate calibration and validation of new functional assays and computational prediction tools.

## Conclusion

These data illustrate the utility and flexibility of the PS4-likelihood ratio methodology in conjunction with the enhanced power afforded from integrating large enriched clinical datasets with unselected case-control datasets. This enabled analysis of 13,966 rare variants in total, including 10,817 missense variants, generating evidence for classification for 6,434 variants overall and 4,690 missense variants across five breast cancer predisposition genes widely tested clinically. We illustrate generation of case-control evidence towards classification of benignity, which is integral to the forthcoming v4.0 revision to the ClinGen ACMG variant classification framework (having previously only allowed case-control evidence towards pathogenicity). We demonstrate the versatility of this tool for quantitative assessment of association against different target odds ratios, allowing evaluation of variants in both high penetrance genes and moderate penetrance genes, as well as for identification of variants of atypical penetrance. These approaches will become increasingly important as there is increasing recognition within variant interpretation of the importance of accounting for variable penetrance.

## Declarations

### Ethics approval and consent to participate

NDRS Data used in these analyses are collected and stored securely within NDRS under the legal permissions afforded by Section 254 of the UK Health and Social Care Act 2012, which permits the collection of patient data without requiring informed consent. This research has been conducted using the UK Biobank Resource under Application Number 76689.

### Consent for publication

Not applicable

### Availability of data and materials

The dataset(s) supporting the conclusions of this article is(are) included within the article (and its additional file(s)). UK Biobank data used in these analyses are available from UK Biobank upon request and approval of a project application (https://www.ukbiobank.ac.uk). Summary data from the BRIDGES consortium is available at the following link: https://www.ccge.medschl.cam.ac.uk/breast-cancer-association-consortium-bcac/data-data-access/summary-results/bridges-summary-results. Data access for CARRIERS, NDRS, and Ambry data was attained on request and is subject to application and project approval from the respective team leads. All other datasets generated or analysed during this study are included in this published article and its supplementary information files. Scripts used to generate the results in this manuscript are available at https://github.com/instituteofcancerresearch/PS4_IntegratingEnrichedData.

## Funding

S.A. and C.F.R. are supported by CG-MAVE, CRUK Programme Award [EDDPGM-Nov22/100004]. For the purpose of Open Access, the author has applied a CC BY public copyright licence to any Author Accepted Manuscript (AAM) version arising from this submission. **BRIDGES:** BCAC is funded by Cancer Research UK [C1287/A16563], the European Union’s Horizon 2020 Research and Innovation Programme (grant numbers 634935 and 633784 for BRIDGES and B-CAST respectively).The BRIDGES panel sequencing was supported by the European Union Horizon 2020 research and innovation program BRIDGES (grant number, 634935) and the Wellcome Trust (v203477/Z/16/Z). All studies and funders are listed in Breast Cancer Association Consortium et al. (N Engl J Med. 2021). **CARRIERS:** Supported in part by the National Institutes of Health (NIH) (grants R01CA192393, R01CA225662, and R35CA253187), the NIH Specialized Program of Research Excellence (SPORE) in Breast Cancer (grant P50CA116201), and the Breast Cancer Research Foundation. All studies and funders are listed in Hu et al. (N Engl J Med. 2021).

### Author’ contributions

Funding acquisition: C.T.; Conceptualisation-case control integration: C.T., M.J., S.A., C.R., Conceptualisation - PS4-LR approach: C.T., C.F.R., C.L., M.J., S.A.; Submission of raw data to NDRS: CanVIG-UK; Data Curation: F.C., M.R., T.P., J.P., K.L., F.M., S.V., S.H., B.T., L.L.; Formal Analysis: S.A., C.F.R.; Investigation: S.A.; Methodology - development of PS4-LR-Calculator framework: C.F.R., C.L., S.A., M.J., C.T.; Methodology – enrichment factor calculation and combination of datasets: C.T., S.A., C.F.R., A.G.; Methodology - update to derivation and boundary specification: C.L., C.F.R., S.A.; Project Administration: S.A., Supervision: C.T.; Visualisation: S.A.; Validation – methodological review: C.T., C.F.R., S.A., M.J., C.L.; Validation - verification of PS4-LR-Calculator framework in Lean4: C.L.; Validation – clinical data application approach: R.A., M.D., G.J.B., A.C., R.R., J.F., B.F., S.P.S., J.G., J.P., T.McD., K.S., H.H., T.McV., CanVIG-UK; Writing – original draft: C.T., S.A.; Writing - review and editing: all authors.

## Supporting information

Supplementary Tables (Large Excel)

## Data Availability

The dataset(s) supporting the conclusions of this article is(are) included within the article (and its additional file(s)). UK Biobank data used in these analyses are available from UK Biobank upon request and approval of a project application (https://www.ukbiobank.ac.uk). Summary data from the BRIDGES consortium is available at the following link: https://www.ccge.medschl.cam.ac.uk/breast-cancer-association-consortium-bcac/data-data-access/summary-results/bridges-summary-results. Data access for CARRIERS, NDRS, and Ambry data was attained on request and is subject to application and project approval from the respective team leads. All other datasets generated or analysed during this study are included in this published article and its supplementary information files.

## Acknowledgements

This work uses data that has been provided by patients and collected by the NHS as part of their care and support. The data are collated, maintained and quality assured by the National Disease Registration Service, which is part of NHS England.

Members of the CanVIG-UK Consortium are listed as followed: Clare Turnbull, Alice Garrett, Lucy Loong, Beth Torr, Charlie F Rowlands, Sophie Allen, Zeid Kuzbari, Miranda Durkie, George J. Burghel, Rachel Robinson, Alison Callaway, Joanne Field, Bethan Frugtniet, Sheila Palmer-Smith, Jonathan Grant, Judith Pagan, Trudi McDevitt, Lowri Hughes, Elizabeth Johnston, Laura Yarram-Smith, Peter Logan, Laura Reed, Katie Snape, Terri McVeigh, Helen Hanson, Marc Tischkowitz, D. Gareth Evans, Fiona Lalloo, Shalaka Samant, Alex Murray, Angela Brady, Ramsay Bowden, Charlene Crosby, Charlotte Jenkins, Christopher Bowles, Dawn O’Sullivan, Deirdre Donnelly, Dhara Randhawa, Emer Atkinson, Elizabeth Johnston, Fiona McRonald, Gillian Rea, Helen Williamson, Helena Calrey, James Tellez, Jennet Williams, Joanna Campbell, Joanna Tolmie, Joanne Mason, John Burn, Jonathan Callaway, Kevin Monahan, Kai Ren Ong, Karen Cadoo, Kezia Quigley, Kristina Stone, Lorainne Hawkes, Louise Izatt, Louise Kiely, Marion Bartlett, Martina Owens, Megan Carney, Miriam J Smith, Munaza Ahmed, Nicola Roberts, Odette Middleton, Philip Dean, Philippa May, Rachel Harrison, Rachel Hart, Ruth Cleaver, Sabrina Talukdar, Samantha Ann Butler, Sean Hegarty, Tina Bedenham, Treena Cranston, Zoe Kemp, Clare Corbett, Esta Cross, Frances Ryan, Helen Lindsay, Jennie Dring, Sarah Shepherd, Zoe Allen, Rowena Mitchell, Inderjit Doal, Florentina Sava, Ciaran McCarthy, Ramanand Jeeneea, Ann-Britt Jones, Maureen Ramos, Donna Lobo, Jessica Gabriel, Bex Pleasance, Dushy Navarajasegaran, Anthony Bench, Patrick Wightman, Joshua Bott, Sally Thomas, Chloe Cassidy, Nina Orfali, Mary Edis, Phil Ostrowski, Aoife Dunne, Laura Freestone, Caroline Shak, Shona Borland, Jessica Willan, Oluwatosin Taiwo, Julita Jaudzemaite, Rubayet Sharmin, Louise Young, Eleanor Mortensson, Rachel Moore, Elisabeth Rolf, Daniel Wood, Jessie Sims, Ross Heron, Laura Elmhirst, Lampros Mavrogiannis, Lihui Wang, Amal Singh, Rachel Salmon, Georgia Thodi, Sin I Chu, Mohamed Ukash, Victoria Steventon-Jones, Ciara Batterton, Gemma Baker, Leigh Jackson, Martin Slater, Kate Annesley, Andrea Western, Eleanor Phillips, Ellen Higgs, Matthew Gordon, Riyaad Aungraheeta, Rachel Price, Shaun Prior, Luke Redford, Amy Clarkson, Navodya Gamage, Sally Halsall, Robert Howie, Ryan Jeavons, Katrina Newland, Diana Walsh, Carryl Dryden, Emily McDermott, Mary Alikian, Lara Hawkes, Lucy Eastoe, Matthew Smith, Sunil Bagha, Victoria Malone, Lauren Silcock, Abbie Dell, Josh Grant, Ivan Oshea, Michael Coghlan, Jennifer Brisbane, Emily Jones, Rebecca Bastock, Diana Rios Szwed, Elizabeth Ratsma, Martyna Borak, Ghada Afifi, Laura Margetts, Lindsey Vialard, Nishath Hamza.

