## Supplementary Material for "Integrating enriched case data from national laboratory testing with population-based case-control analyses: a novel statistical likelihood-ratio methodology for PS4 applied to 325,345 breast cancer cases and 671,006 controls"

^2^ NHS England, National Disease Registration Service, United Kingdom

^3^ St George's University Hospitals NHS Foundation Trust, Tooting London, United Kingdom

^4^ Department of Laboratory Medicine and Pathology, Mayo Clinic, Rochester Minnesota, USA

^5^ Ambry Genetics, Aliso Viejo California, USA

^6^ Sheffield Children's NHS Foundation Trust, Sheffield , United Kingdom

^7^ Manchester University Hospitals NHS Foundation Trust, Manchester , United Kingdom

^8^ The University of Manchester, Manchester, United Kingdom

^9^ University Hospital Southampton NHS Foundation Trust, Salisbury, United Kingdom

^10^ Leeds Teaching Hospitals NHS Trust, Leeds , United Kingdom

^11^ Nottingham University Hospitals NHS Trust, Nottingham , United Kingdom

^12^ Cardiff and Vale University Health Board, Cardiff , United Kingdom

^13^ NHS Greater Glasgow and Clyde, Glasgow , United Kingdom

^14^ Western General Hospital, Edinburgh, United Kingdom

^15^ CHI at Crumlin, Dublin, Ireland

^16^ Royal Devon University Healthcare NHS Foundation Trust, Exeter, United Kingdom

^17^ University of Exeter, Exeter, United Kingdom

^18^ The Royal Marsden NHS Foundation Trust, London, United Kingdom

^19^ Overdog.ai Ltd, London, United Kingdom

### **Supplementary Methods**

#### Dataset Curation

For all datasets, variants were annotated according to the build 37 (GrCh37) versions of MANE select transcripts for each gene (*BRCA1*: NM_007294.3/ENST00000357654.3, *BRCA2*: NM_000059.3/ENST00000544455.1, *PALB2*: NM_024675.3/ENST00000261584.4, *ATM*: NM_000051.3/ENST00000278616.4, *CHEK2*: NM_007194.3/ENST00000328354.6).

##### UK Biobank (UKB)

Per-patient whole exome sequencing data was accessed on 20/08/2025^1^. In total, 18,477 female individuals with reported personal history of breast cancer (*in situ* or invasive, any grade or histological subtype), either self-reported or from linkage to national cancer registrations (cases), and all 419,373 individuals with no reported history of breast cancer (controls) were included. Data was curated as defined previously^2^, with an additional filter to restrict to “White” ancestry (top-level self-reported parent ancestry, individuals associated with multiple ethnicities or “Other” removed).

##### CARRIERS Consortium

Per-patient summary-level data from 32,247 female patients with breast cancer (*in situ* or invasive) and 32,544 female controls from across 12 studies, filtered for “Non-Hispanic White” individuals. Methods for collection as previously described.^3^

##### BRIDGES Consortium

Per-variant summary-level data from 42,062 female patients with breast cancer (*in situ* or invasive) and 44,035 female controls from across 30 BCAC (Breast Cancer Association Consortium) studies which recruited patients unselected for family history, and further filtered for individuals of European ancestry (<https://www.ccge.medschl.cam.ac.uk/breast-cancer-association-consortium-bcac/data-data-access/summary-results/bridges-summary-results>). Methods for collection of data as previously described.^4^

##### UK National Germline Data (NDRS)

###### Genetic testing data

Diagnostic and predictive germline testing data for *BRCA1* and *BRCA2* were and continue to be submitted by 17 NHS genetic testing laboratories centrally to the National Disease Registration Service (NDRS). Laboratories involved in submission are represented within the Cancer Variant Interpretation Group UK (CanVIG-UK). Variant submission was largely of pathogenic and likely pathogenic variants, with some laboratories also submitting VUS. *PALB2*, *ATM*, and *CHEK2* variants submitted from UK laboratories were excluded from this analysis due to incomplete laboratory submissions for these genes, in particular due to UK clinical testing and reporting for *ATM* and *CHEK2* focusing on protein truncating variants rather than missense variants.

Germline data collection was censored at 24/07/2024, with germline data available for some laboratories from 2000-2024. Data fields included the gene, coding DNA change, predicted protein impact, variant classification (where provided), and total count in full-screen/diagnostic tests (where one observation = one detection in an independently ascertained family). Variant nomenclature was verified using VariantValidator^5^. Variants where the gene and variant nomenclature did not match following the VariantValidator check, and the true variant could not be confidently identified, were removed from analysis.

###### Eligibility for genetic testing

In 2018, the National Genomic Test Directory for Rare and Inherited Disease was released.^6^ The directory included a set of eligibility criteria for offering genetic testing, including for patients with breast cancer under eligibility criteria R208. These criteria have changed over time (Box S1):

Living affected individual (proband) with breast cancer where the individual +/- family history meets one of the criteria. The proband has:

a. Breast cancer (age <40 years), OR
b. Bilateral breast cancer (age < 60 years), OR
c. Triple negative breast cancer (age < 60 years), OR
d. Assigned male at birth and affected with breast cancer (any age), OR
e. Breast cancer (age <45 years) and a first degree relative with breast cancer (age <45 years), OR
f. Combined pathology-adjusted Manchester score ≥15 or single gene pathology adjusted score of ≥10 or BOADICEA/CanRisk score ≥10% OR
g. Ashkenazi Jewish ancestry and breast cancer at any age
h. ≥ 1 grandparent from Westray (Orkney) or Whalsay (Shetland) and breast cancer at any age

**Box S1:** Current eligibility criteria for R208: Inherited breast cancer for living patients with breast cancer, as per the National Genomic Test Directory for Rare and Inherited Disease, v8.1, July 2025

###### Cancer Registry

Individuals with breast cancer in England were identified using the National Cancer Registration Dataset (NCRD) which is managed by the National Cancer Registration and Analysis Service (NCRAS) and collects data on all cancer diagnoses in England.^7^ Breast cancer was defined as malignant neoplasm of the breast (ICD-10 C50), excluding stage 0 cases (Paget’s disease), and/or intraductal carcinoma in situ of breast (ICD-10 D05.1) provided these were high-grade cases (GH, G3, or G4). Other data collected included self-identified ethnicity and gender.

Cancer registry data from NCRAS was linked to the germline genetic testing dataset using pseudonymised identifiers based on NHS number, Date of Birth, and Postcode to identify 44,917 white, female breast cancer patients tested for *BRCA1* or *BRCA2* variants. Patients with a *BRCA1* or *BRCA2* variant who did not link to the cancer registry, had an ethnicity which was not “White”, or had a gender other than Female, were excluded from analysis.

##### Ambry Genetics (Ambry)

Diagnostic testing results from Ambry were censored at 25/03/2025. Per-patient summary-level data was collated from 187,642 self-reported “White”/“Caucasian” female patients with breast cancer, tested by Ambry Genetics for *BRCA1, BRCA2, PALB2, ATM* or *CHEK2* variants between 2015-2024. Variant nomenclature was verified using VariantValidator^5^. Variants where the gene and variant nomenclature did not match following the VariantValidator check, and the true variant could not be confidently identified, were removed from analysis. Liftover from build 38 to build 37 was performed for all variants to align with chromosome build used for all other datasets.

Breast cancer was defined as malignant neoplasm of the breast (ICD-10 C50). Other data included self-reported ethnicity, age at testing, sex, year tested, test name and code, gene, RefSeq transcript, coding DNA change, predicted protein impact, zygosity, classification, and the following information provided as binary ‘yes/no’ responses based on ICD-10 code: personal history of breast cancer, personal history of ovarian cancer, family history of breast cancer, family history of ovarian cancer. Patients were excluded if they had a self-reported ethnicity other than “White”/”Caucasian”, and/or were tested for a single site or single gene, as these patients may have been identified through cascade testing.

##### gnomAD v4.1.0

###### Data used to identify variants eligible for BA1

gnomAD v4.1.0 joint frequency VCF files were downloaded from the gnomAD website on 19/03/2025^8^. Variants with a filter of “PASS”, “EXOMES_FILTERED”, and “GENOMES_FILTERED” were included. VCF files were filtered for each gene using the following genomic coordinate ranges, per regions provided by gnomAD: *BRCA1*, 17:43044295-43170245; *BRCA2*, 13:32315086-32400268; *PALB2*, 16:23603160-23641310; *ATM*, 11:108223044-108369102; *CHEK2*, 22:28687742-28742422. The following fields were extracted from the VCF: #CHROM, POS, ID, REF, ALT, FILTER, faf95_max_joint, faf95_max_genomes, faf95_max_exomes

GroupMAX 95% filtering allele frequency (FAF) across all non-bottlenecked genetic ancestry groups (not including Amish, Ashkenazi Jewish, European Finnish, and Remaining Individuals) were extracted for all variants. The GroupMax FAF was compared to the maximum tolerated allele frequency (MTAF) for each gene, as defined by the respective gene variant curation expert group (VCEP); if the GroupMax FAF was higher than the gene MTAF, the variant was considered to have met stand-alone evidence criteria for benignity (BA1) and was added to a “BA1 list”. Liftover from build 38 to build 37 was performed for all variants in the BA1 list using Ensembl Assembly Converter (<https://mart.ensembl.org/Homo_sapiens/Tools/AssemblyConverter>). All variant nomenclature was validated using VariantValidator^5^.

###### Data used as population controls for case-control analysis

gnomAD v4.1.0 exome data VCF files were downloaded from the gnomAD website on 19/03/2025 (<https://gnomad.broadinstitute.org/data>). Exome data files were chosen for this purpose as these files provide separate non-UKB counts, which avoids potential double-counting of controls with the UKB dataset. VCF files were filtered for relevant gene regions as above for the joint frequency data. Variants in these regions were further filtered to require QC filter = “PASS”.

The following fields were extracted from the VCF: #CHROM, POS, ID, REF, ALT, FILTER, AC_non_ukb_nfe, AN_non_ukb_nfe, nhomalt_non_ukb_nfe, and faf95_non_ukb_nfe. Allele counts were converted to individuals by removing n=1 count for each homozygous carrier instance, and allele number halved. Liftover from build 38 to build 37 was performed for all variants in the BA1 list using Ensembl Assembly Converter (<https://mart.ensembl.org/Homo_sapiens/Tools/AssemblyConverter>). All variant nomenclature was validated using VariantValidator^5^.

#### PS4-LR-Calc methodology

Further to the methodology described in Rowlands et al. (2024)^9^, for the analyses presented in the main text, we have incorporated an updated approach for the establishment of hypothetical odds-to-probability mapping. For the following distribution of case and control variant observations:

|  | Cases | Controls |
| --- | --- | --- |
| Carriers | $a$ | $b$ |
| Non-carriers | $c$ | $d$ |

The probability of case selection among carriers generating a target odds ratio of interest of $x$ was previously defined as:

$$p\left( OR=x \right)=\frac{xc}{xc+d}$$

To scale this probability according to any target odds ratio, Under the updated model, we first construct a table of expected pseudocounts under probability of case selection among carriers $p$:

|  | Cases | Controls |
| --- | --- | --- |
| Carriers | $(a+b)\cdot p$ | $(a+b)\cdot(1-p)$ |
| Non-carriers | $a+c-\left( a+b \right)\cdot p$ | $b+d-(a+b)\cdot(1-p)$ |

The resulting odds ratio $x$ under probability of case selection among carriers $p$ is given by the following equation:

$$x=\frac{(a+b)\cdot p}{a+c-\left( a+b \right)\cdot p}\cdot\frac{b+d-(a+b)\cdot(1-p)}{(a+b)\cdot(1-p)}$$

Which simplifies to:

$$x=\frac{p}{a+c-\left( a+b \right)\cdot p}\cdot\frac{b+d-(a+b)\cdot(1-p)}{1-p}$$

Through further simplification and rearrangement, it can be shown that, for a given set of variant counts, the probability of case selection among carriers $p$ and odds ratio $x$ satisfy the following quadratic equation:

$$\left( a+b \right)\left( x-1 \right)p^{2}-\left( x\left( 2a+b+c \right)+(d-a) \right)p+x\left( a+c \right)=0$$

Where the target OR equals 1 (i.e. $x=1$), the quadratic term equals 0 and the equation simplifies to:

$$a+c-(2a+b+c+d-a)p=0$$

$$\therefore p=\frac{a+c}{a+b+c+d}$$

i.e. $p$ equals the proportion of all samples in the dataset which belong to the case series.

For all other target odds ratios, the corresponding probability of case selection among carriers are instead calculated using the quadratic formula as follows:

$$p\left( OR=x \right)=\frac{x\left( 2a+b+c \right)+(d-a)-\sqrt{{(x\left( 2a+b+c \right)+(d-a))}^{2}-4x(a+b)(x-1)(a+c)}}{2(a+b)(x+1)}$$

These updated equations were used to generate the data presented in the main text. This adjustment provides refined estimates of the probability generating an odds ratio of interest. This refinement is of negligible impact when case and control numbers are balanced as per scenarios presented, but will impact the LR in scenarios in which there is very substantial imbalance between the number of cases and number of controls.

#### Dataset combination

Protein-truncating variants (PTVs) were defined as variants with one of the following variant effect consequences, annotated using Ensembl VEP^10^ (v112): "stop_gained", "splice_acceptor_variant", "splice_donor_variant", "frameshift_variant". PTVs that were in the final exon of a gene and/or splice variants that were excluded from primary analysis of the BRIDGES dataset^4^ were removed. Missense variants were defined as variants with a VEP consequence of “missense_variant”. All variant nomenclature was verified using VariantValidator^5,11^.

All datasets were merged on gene name and coding DNA nomenclature. On creation of the combined dataset, the dataset was additionally merged with the list of BA1-eligible variants. Any variant present in the BA1-eligible variant list was removed.

The likelihood ratio cannot be modelled by the binomial distribution underlying the PS4 Likelihood Ratio Calculator (PS4-LR-Calc) where carrier counts are too large. 10 variants in the Ambry dataset had a case and/or control carrier count exceeding these limits while still remaining ineligible for BA1, and therefore PS4-LLR could not be calculated. For these 10 variants, the Ambry dataset was not included in the dataset combination (retaining all data from other datasets where available), and variants for which this is applicable have been annotated in Supplementary Table 2.

#### Per-variant case-control analysis

###### Single dataset

Likelihood Ratio (LR) was calculated using the PS4-LR-Calc as published^9^ using a default target odds of association of 4 for high-penetrance genes (*BRCA1*, *BRCA2*, *PALB2*), and 2 for moderate penetrance genes (*ATM*, *CHEK2*), as per published recommendations for high increased risk and moderate increased risk for breast cancer relative to the general population risk^12,13^.

Fisher’s exact test of independence and quantification of effect size using an odds ratio (OR) were used to test and measure the difference in carrier frequency between variant heterozygotes with breast cancer versus breast cancer unaffected individuals. The OR and 95% confidence intervals (lower 95% confidence interval = LCI, upper 95% confidence interval = UCI) were calculated as described above. Where variant carriers were only identified in controls, OR and respective confidence intervals are not calculated and are set to 0. Where variant carriers were only identified in cases, the Haldane-Anscombe correction was applied (+0.5 to each value) to enable OR calculation.

###### Unselected dataset meta-analysis

A meta-analysis was conducted using methodology as previously described^2^. Briefly, data from the three unselected datasets (BRIDGES, CARRIERS, UKB) was used to calculate per-variant betas of association with breast cancer, OR, and standard error estimates with upper and lower 95% confidence intervals for each estimate where case data was available for at least two datasets. Meta-analysis was conducted using a fixed effect inverse-variance approach, unadjusted for age or other metrics from the constituent studies. *p_-_*values were derived from a Fisher exact test where there were only two contributing datasets and at least one of the carrier counts was <6; otherwise, *p*-values were derived from a chi-squared test.

Where variant carriers were observed in cases only, the Haldane correction was applied (+0.5 to each value) to enable calculation of an OR. Where variant carriers were observed in controls only, data was not used to calculate an OR and not included in the meta-analysis to avoid generation of effect size where the variant was not seen in cases.

###### Combined dataset

For each variant, data available in each dataset was assessed for inclusion against the delineated rules (Supplementary Table 14). After removing data which did not meet these rules, the LR from each dataset was combined as follows:

$${LR}_{tot}= {LR}_{1}\times{LR}_{2}\times{LR}_{3}\ldots\times{LR}_{n}$$

using data from $n$ datasets for which an LR could be calculated. Where the combined likelihood was too small for storage within R systems, LR was set to the smallest stored value possible (5.0x10^-324^) The total amount of evidence points (PS4-LLR) was re-calculated for LR_tot_ by way of log transformation (base 2.08), as per the Tavtigian et al. scaled point systems^14^:

$$PS4\_LLR= {{log}_{2.08}(LR}_{tot})$$

Comparison of data for each gene was undertaken using the Kruskal-Wallis rank sum test, and the Dunn test to identify which genes had statistically different distributions. To account for multiple testing, the Bonferroni-adjusted p-value for significance was set at 0.005.

#### Reduced penetrance criteria

Four criteria were defined to identify potentially reduced penetrance variants when comparing standard (OR≥4) to reduced penetrance (OR≥2) target odds of association. For any criteria to apply, a minimum change of two evidence strength categories was required.

- Evidence strength changes direction (Benign to Pathogenic)
- Loses benign evidence (Benign Moderate, Strong, or Very strong to No Evidence)
- Gains pathogenic evidence (No evidence to Pathogenic Moderate, Strong, or Very Strong)
- Substantially strengthens weakly pathogenic evidence (Pathogenic Moderate to Very Strong, or Pathogenic Supporting to Strong or Very Strong)

All criteria were defined based on the impact on an overall classification for a reduced penetrance variant in providing or strengthening evidence available towards pathogenicity. Where benign evidence was applicable at a standard target odds≥4 but was removed at the reduced target odds≥2 (criteria 2), use of a reduced target odds of association removes a potentially contradictory piece of evidence and thus better enables classification as pathogenic for a reduced penetrance variant.

#### Reclassification of missense VUS

As per the guidelines recommended by the ClinGen ENIGMA *BRCA1* and *BRCA2* Variant Curation Expert Panel (VCEP)^12^, variants with results from high-throughput functional assays attained either strong evidence for PS3 or BS3 as assigned by the VCEP Specifications Table 9 (Supplementary Table 15)^15-25^. Variants in potentially clinically important functional domains (*BRCA1*: p.2-101, p.1391-1424, and p.1650-1857; *BRCA2*: p.10-40, p.2481-3186) with data from BayesDel^26^ and SpliceAI^27^ exceeding VCEP-recommended thresholds attained supporting evidence for PP3 or BP4 variants, thresholds detailed as follows (PP3: *BRCA1*: BayesDel no-AF score ≥0.28 OR SpliceAI ≥ 0.2 at any position; *BRCA2*: BayesDel no-AF score ≥0.30 OR SpliceAI ≥ 0.2 at any position. BP4: *BRCA1*: BayesDel no-AF score ≤0.15 AND SpliceAI ≤ 0.1; *BRCA2*: BayesDel no-AF score ≤0.18 AND SpliceAI ≤ 0.1).

### **Supplementary Figures**

Supplementary Figure 1: Boxplot of PS4-LLR evidence assigned for 1,744 rare PTVs included in the combined dataset. Each dot represents a single variant. Red dashed line represents 0 evidence points attained. 11 variants with PS4-LLR higher than +160 were set at +160, 1 variant with PS4-LLR less than -160 was set at -160. Statistically significant differences between average LLR for each gene are displayed (Kruskal-Wallis rank sum: p-value=1.82x10^-15^, Bonferroni-adjusted p-value threshold for significance=0.005)

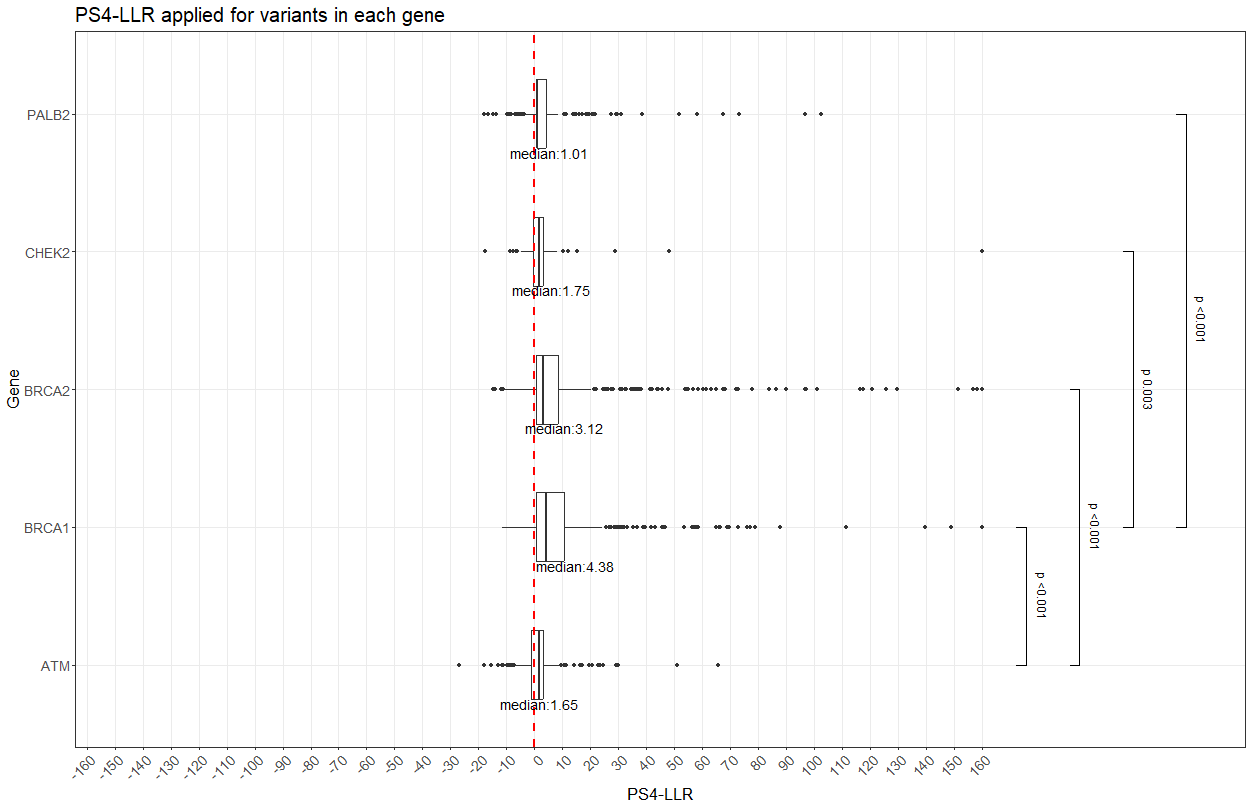

Supplementary Figure 2: Case-control LLR for variants classified as pathogenic, likely pathogenic, likely benign, or benign in ClinVar with at least 2 star classifications for each gene. A: *BRCA1* (Total number of variants (n) = 146). B: *BRCA2* (n=126). C: *PALB2* (n=2). D: *ATM* (n=33). E: *CHEK2* (n=5).

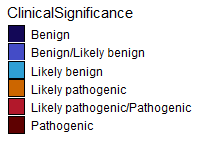

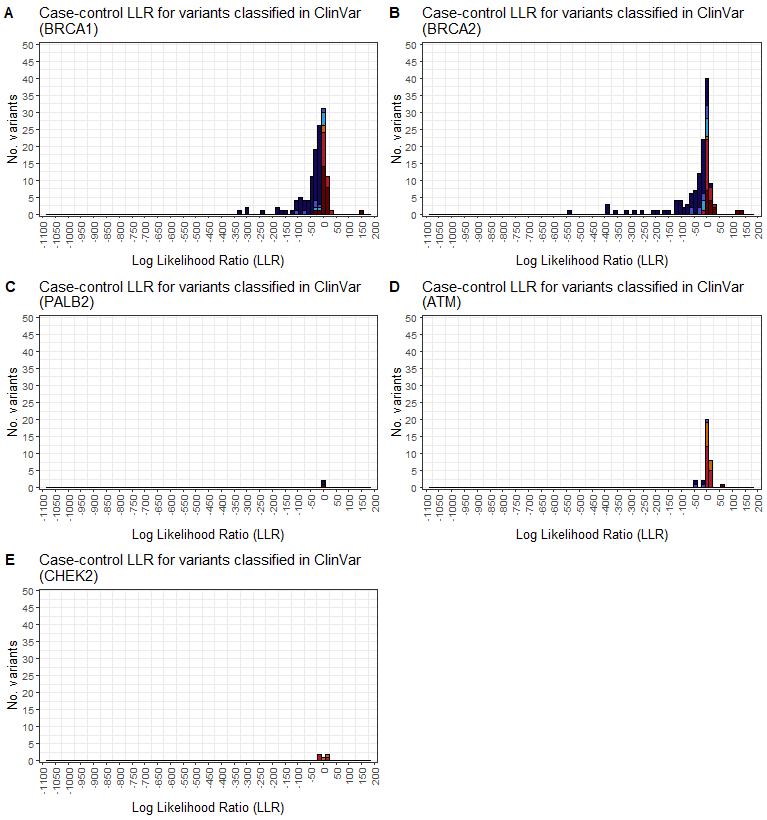

#### Supplementary Figure 3: Sankey diagrams demonstrating difference of evidence application for rare missense variants at a target odds of association of OR≥2 and OR≥4. A: BRCA1; B: BRCA2; C: PALB2; D: ATM; E: CHEK2. P_VSTR=Pathogenic Very Strong; P_STR=Pathogenic Strong; P_MOD=Pathogenic Moderate; P_SUP=Pathogenic Supporting; B_VSTR=Benign Very Strong; B_STR=Benign Strong; B_MOD=Benign Moderate; B_SUP=Benign Supporting.

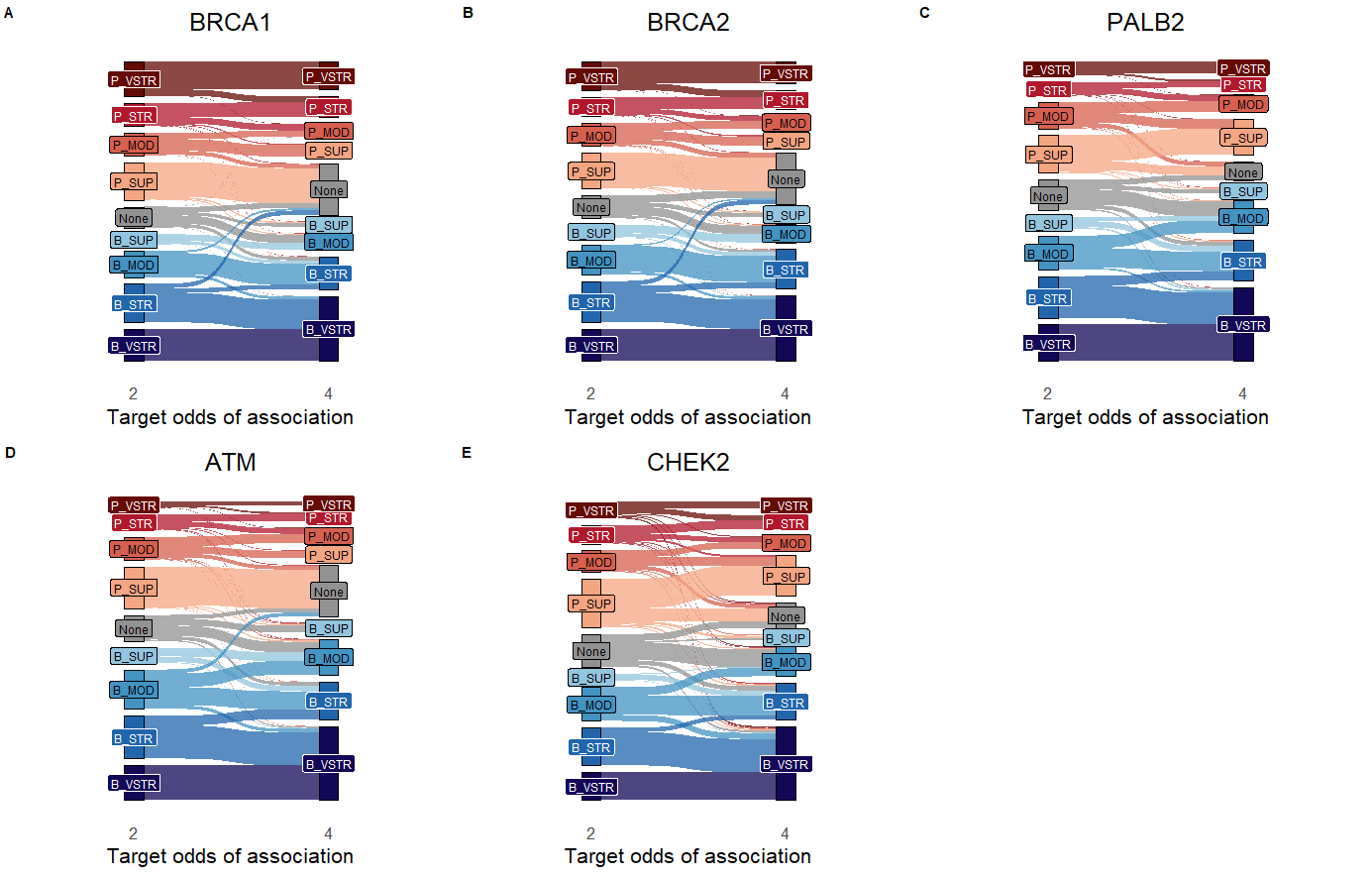

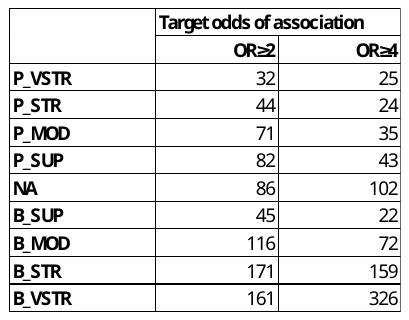

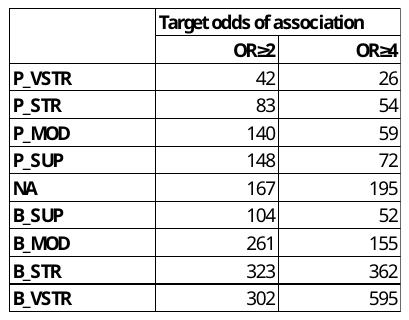

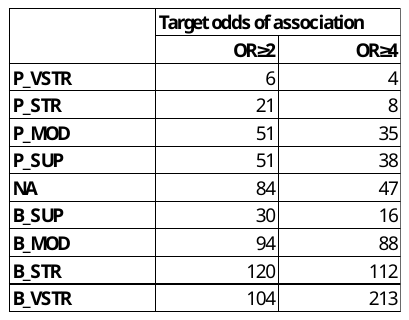

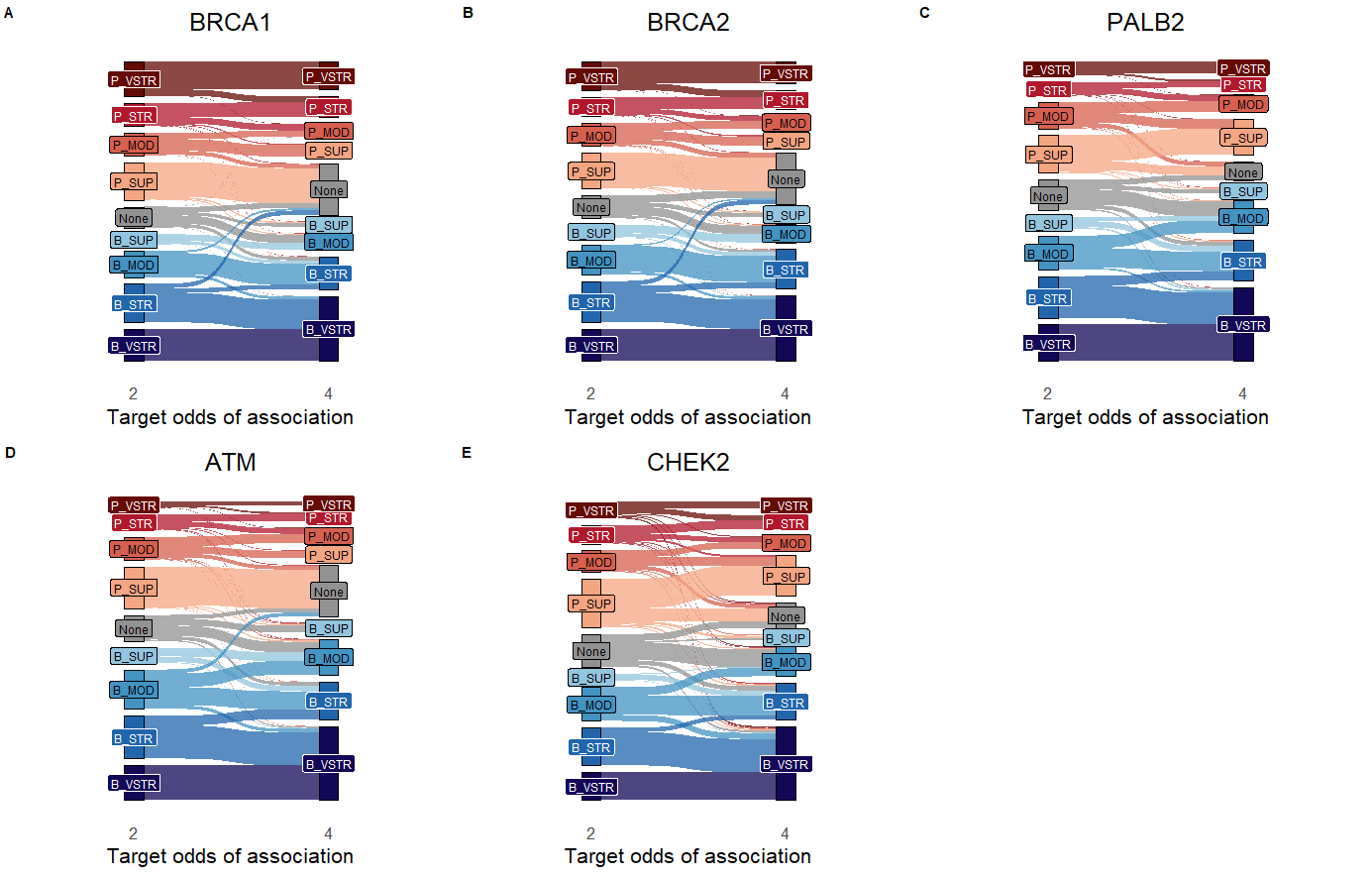

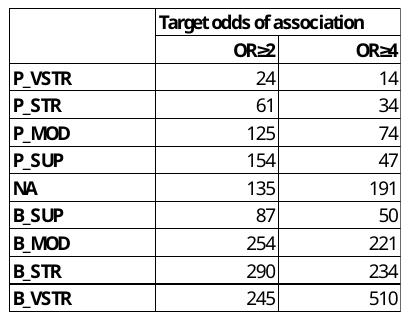

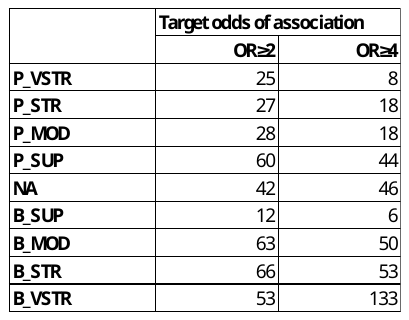

Supplementary Figure 4: Case:control ratio for NDRS and UKB case series with varying proportions of 419,373 controls. The point at which case:control ratio is equal for both datasets is at a ratio of 1 case:6.62 controls (297,141 controls assigned to NDRS cases, and 122,232 controls assigned to UKB cases).

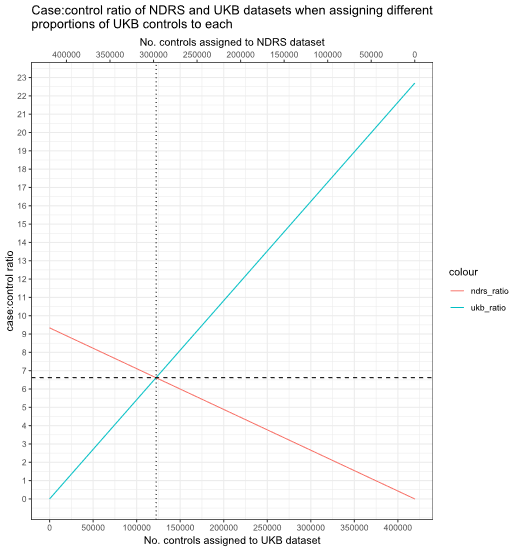

Supplementary Figure 5: ACMG points applicable for 1, 2, or 3 observations in cases or controls at a target odds of association of OR≥4 for case:control ratios ranging from 0-12. A-C: Total controls exceed cases in a 10:1 ratio, with 0 case carriers, and either A: 1 control carrier; B: 2 control carriers; C: 3 control carriers. D-F: Total cases exceed controls in a 10:1 ratio, with 0 control carriers, and either D: 1 case carrier; E: 2 case carriers; F: 3 case carriers.

The size imbalance at which 0 ACMG points are applicable for each scenario is plotted as a solid black line. A ratio larger than this will produce “false positive” (when total controls exceed cases) or “false negative” (when total cases exceed controls) results for the respective observations of the variant. The number of ACMG points applicable for a dataset with a size discrepancy of 10x is plotted in red to demonstrate this concept; the size discrepancy produces false positive results in A and B, but not C.

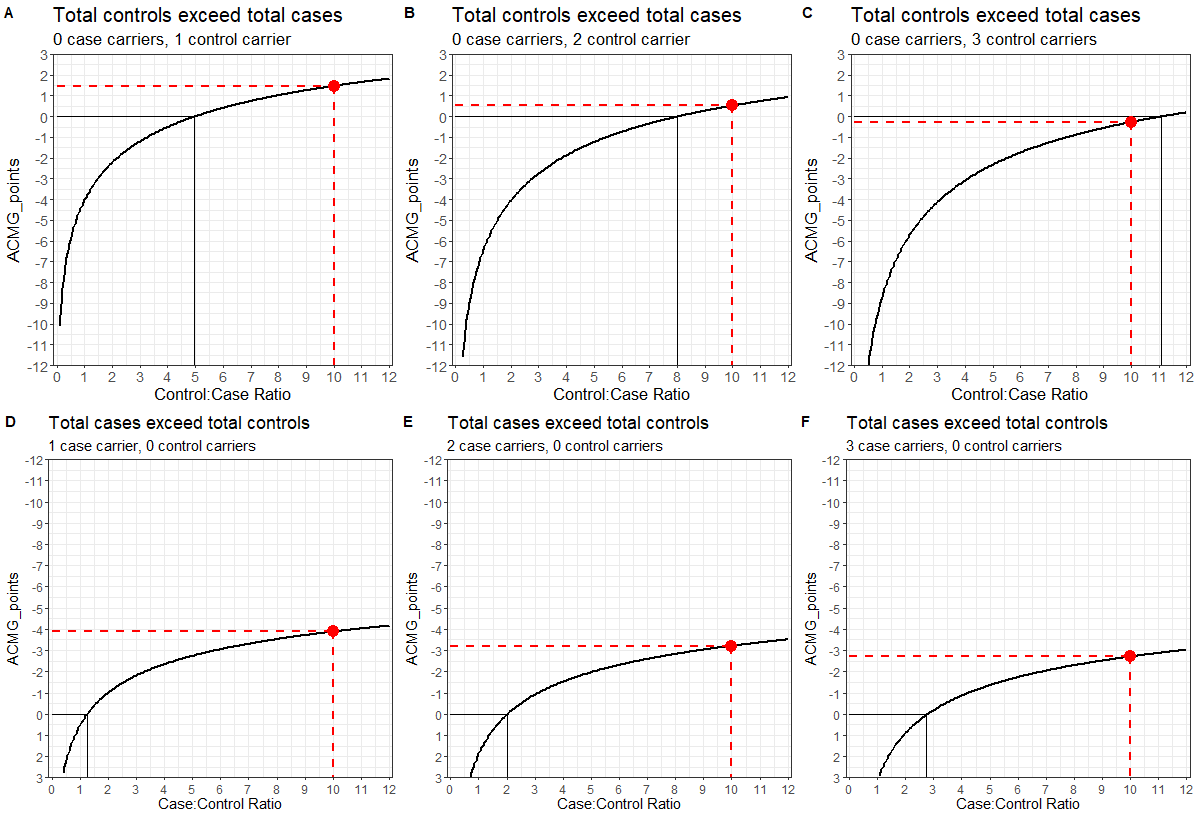

Supplementary Figure 6: Observed OR plotted against likelihood space for all combinations of 0-100 cases and 0-100 controls. OR≥Observed Odds Ratio; LR_percent=Percentage of the binomial distribution covered by likelihood space; CI=95% confidence interval. A target odds of association of OR≥4 and target odds of non-association of OR≤1 was used. A case cohort denominator of 42,062 and control cohort denominator of 44,035 was used for each calculation. A: Plot of all case and control combinations. B: Plot of all case and control combinations which have an upper CI below 4 and a lower CI above 1.

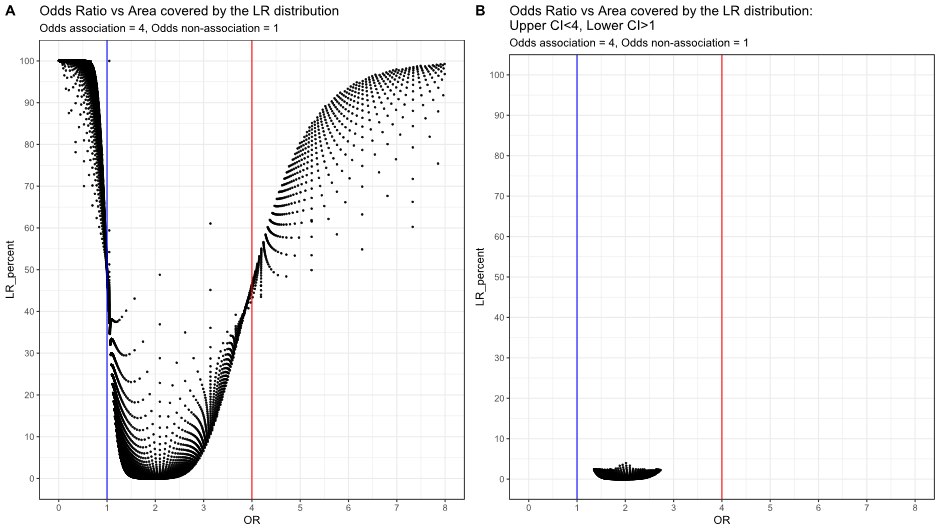

### **Supplementary Tables**

Supplementary Table 1: Allele frequency of PTVs in each constituent dataset (unselected and enriched). EF=Enrichment Factor; CI=Confidence Interval.

|  | | **Unselected breast cancer datasets** | | | | **NDRS** | | | | **Ambry** | | | |
| --- | --- | --- | --- | --- | --- | --- | --- | --- | --- | --- | --- | --- | --- |
|  |  | **No. cases with PTVs** | **Total tested** | **Allele Frequency** | **Average Allele Frequency** | **No. cases with PTVs** | **Total tested** | **Allele Frequency** | **EF (95% CI)** | **No. cases with PTVs** | **Total tested** | **Allele Frequency** | **EF (95% CI)** |
| ***BRCA1*** | **UKB** | 94 | 18477 | 0.005 | 0.007 | 1606 | 44917 | 0.036 | 4.84  (4.43-5.29) | 3642 | 187642 | 0.019 | 2.63  (2.42-2.85) |
|  | **BRIDGES** | 427 | 42062 | 0.010 |  |  |  |  |  |  |  |  |  |
|  | **CARRIERS** | 164 | 32247 | 0.005 |  |  |  |  |  |  |  |  |  |
| ***BRCA2*** | **UKB** | 242 | 18477 | 0.013 | 0.012 | 2165 | 44917 | 0.048 | 3.92  (3.65-4.21) | 5688 | 187642 | 0.030 | 2.47  (2.31-2.63) |
|  | **BRIDGES** | 609 | 42062 | 0.014 |  |  |  |  |  |  |  |  |  |
|  | **CARRIERS** | 290 | 32247 | 0.009 |  |  |  |  |  |  |  |  |  |
| ***PALB2*** | **UKB** | 105 | 18477 | 0.006 | 0.005 | NA | NA | NA | NA | 1646 | 187642 | 0.009 | 1.92  (1.73-2.14) |
|  | **BRIDGES** | 236 | 42062 | 0.006 |  |  |  |  |  |  |  |  |  |
|  | **CARRIERS** | 82 | 32247 | 0.003 |  |  |  |  |  |  |  |  |  |
| ***ATM*** | **UKB** | 119 | 18477 | 0.006 | 0.006 | NA | NA | NA | NA | 2186 | 187642 | 0.012 | 2.02  (1.84-2.22) |
|  | **BRIDGES** | 269 | 42062 | 0.006 |  |  |  |  |  |  |  |  |  |
|  | **CARRIERS** | 147 | 32247 | 0.005 |  |  |  |  |  |  |  |  |  |
| ***CHEK2*** | **UKB** | 231 | 18477 | 0.013 | 0.013 | NA | NA | NA | NA | 4539 | 187642 | 0.024 | 1.81  (1.70-1.92) |
|  | **BRIDGES** | 710 | 42062 | 0.017 |  |  |  |  |  |  |  |  |  |
|  | **CARRIERS** | 302 | 32247 | 0.009 |  |  |  |  |  |  |  |  |  |

Supplementary Table 2: All 13,966 rare PTV and missense variants identified, and PS4-LLR evidence applied for each (OR≥4 for *BRCA1*, *BRCA2*, and *PALB2*, and OR≥2 for *ATM* and *CHEK2*). For each dataset, the following information is provided: Case and control carrier and total counts, Allele Frequency (AF) in cases and controls, Odds Ratio (OR) with 95% Confidence Intervals, Fisher's exact p-value, Likelihood Ratio (LR), PS4-LLR evidence, and ACMG Evidence Strength. Please note that due to policies regarding identifiable information, variant carrier counts in cases and controls from the UK Biobank dataset have been redacted. Where n=0 for variant carriers, the Haldane correction was applied before calculation of the observed OR and p-value. PS4-LLR was calculated by log conversion to base 2.08 of the LR. For variants in the Ambry datasets where variant carrier count was too large to calculate PS4-LLR, LR and LLR are listed as ‘NC - not calculable’ (see Supplementary Methods).

[See separate Supplementary Table 2]

#### Supplementary Table 3: Summary of all 3,149 rare PTVs identified for each gene, including variants removed before combined PS4-LLR was calculated. Variants attaining PS4-LLR between 0 and -1 in the NDRS dataset are included in the ‘Benign evidence from NDRS set’ column instead of the ‘No evidence applied’ column, as these variants are not included in the final combined PS4-LLR calculation.

|  | | Rare PTVs included in final dataset (N=1,744) | | | Rare PTVs not included in final dataset (N=1,405) | | | | Total number of rare PTVs in entire dataset (N=3,149) |
| --- | --- | --- | --- | --- | --- | --- | --- | --- | --- |
|  |  | Evidence towards pathogenicity (LLR≥1) | No evidence applied | Evidence towards benignity (LLR≤-1) | Single observation (cases or controls) | Variant only had benign evidence from NDRS set | Discrepant data caused by case:control imbalance | 1≥observed OR≥target odds of association (suspected low penetrance) |  |
| *BRCA1* (OR≥4) | UKB | 26 | 5 | 0 | 53 | 0 | 0 | 0 | 84 |
|  | CARRIERS | 16 | 4 | 7 | 65 | 0 | 0 | 0 | 92 |
|  | BRIDGES | 57 | 3 | 5 | 101 | 0 | 0 | 0 | 166 |
|  | NDRS | 146 | 0 | 0 | 192 | 28 | 0 | 0 | 366 |
|  | AMBRY | 119 | 122 | 37 | 118 | 0 | 0 | 0 | 396 |
|  | Combined | 253 | 103 | 31 | 301 | 17 | 0 | 0 | 705 |
| *BRCA2* (OR≥4) | UKB | 61 | 16 | 11 | 87 | 0 | 0 | 1 | 176 |
|  | CARRIERS | 39 | 7 | 6 | 103 | 0 | 0 | 1 | 156 |
|  | BRIDGES | 69 | 10 | 21 | 224 | 0 | 0 | 1 | 325 |
|  | NDRS | 221 | 1 | 0 | 240 | 80 | 0 | 1 | 543 |
|  | AMBRY | 196 | 229 | 78 | 202 | 0 | 0 | 1 | 706 |
|  | Combined | 394 | 198 | 75 | 486 | 33 | 0 | 1 | 1187 |
| *PALB2* (OR≥4) | UKB | 20 | 6 | 2 | 31 | 0 | 0 | 0 | 59 |
|  | CARRIERS | 8 | 3 | 7 | 34 | 0 | 0 | 0 | 52 |
|  | BRIDGES | 19 | 5 | 8 | 66 | 0 | 0 | 0 | 98 |
|  | AMBRY | 124 | 5 | 29 | 58 | 0 | 0 | 0 | 216 |
|  | Combined | 135 | 11 | 32 | 109 | 0 | 0 | 0 | 287 |
| *ATM* (OR≥2) | UKB | 46 | 10 | 4 | 91 | 0 | 18 | 0 | 169 |
|  | CARRIERS | 16 | 12 | 10 | 81 | 0 | 0 | 0 | 119 |
|  | BRIDGES | 27 | 26 | 13 | 134 | 0 | 0 | 0 | 200 |
|  | AMBRY | 245 | 18 | 92 | 244 | 0 | 0 | 0 | 599 |
|  | Combined | 281 | 25 | 105 | 363 | 0 | 3 | 0 | 777 |
| *CHEK2* (OR≥2) | UKB | 11 | 2 | 1 | 17 | 0 | 5 | 1 | 37 |
|  | CARRIERS | 6 | 3 | 2 | 23 | 0 | 0 | 1 | 35 |
|  | BRIDGES | 10 | 2 | 3 | 35 | 0 | 0 | 1 | 51 |
|  | AMBRY | 61 | 6 | 18 | 78 | 0 | 0 | 1 | 164 |
|  | Combined | 71 | 12 | 18 | 91 | 0 | 0 | 1 | 193 |

#### Supplementary Table 4: Summary of all 10,817 rare missense variants identified for each gene, including variants removed before combined PS4-LLR was calculated. Variants attaining PS4-LLR between 0 and -1 in the NDRS dataset are included in the ‘Benign evidence from NDRS set’ column instead of the ‘No evidence applied’ column, as these variants are not included in the final combined PS4-LLR calculation.

|  | | Rare missense variants included in final dataset (N=4,690) | | | Rare missense variants not included in final dataset (N=6,127) | | | | Total number of rare missense variants in entire dataset (N=10,817) |
| --- | --- | --- | --- | --- | --- | --- | --- | --- | --- |
|  |  | Evidence towards pathogenicity (LLR≥1) | No evidence applied | Evidence towards benignity (LLR≤-1) | Single observation (cases or controls) | Benign evidence from NDRS set | Discrepant data caused by case:control imbalance | 1≥observed OR≥target odds of association (suspected low penetrance) |  |
| *BRCA1* (OR≥4) | UKB | 41 | 75 | 112 | 243 | 0 | 0 | 4 | 475 |
|  | CARRIERS | 21 | 17 | 102 | 186 | 0 | 0 | 4 | 330 |
|  | BRIDGES | 36 | 27 | 153 | 383 | 0 | 0 | 4 | 603 |
|  | NDRS | 48 | 3 | 0 | 54 | 659 | 0 | 4 | 768 |
|  | AMBRY | 67 | 84 | 429 | 699 | 0 | 0 | 4 | 1283 |
|  | Combined | 127 | 102 | 579 | 965 | 189 | 0 | 4 | 1966 |
| *BRCA2* (OR≥4) | UKB | 93 | 150 | 209 | 502 | 0 | 0 | 9 | 963 |
|  | CARRIERS | 44 | 30 | 188 | 428 | 0 | 0 | 9 | 699 |
|  | BRIDGES | 74 | 47 | 277 | 886 | 0 | 0 | 9 | 1293 |
|  | NDRS | 66 | 8 | 0 | 111 | 1255 | 0 | 9 | 1449 |
|  | AMBRY | 108 | 170 | 851 | 1377 | 0 | 0 | 9 | 2515 |
|  | Combined | 211 | 195 | 1164 | 1966 | 330 | 0 | 9 | 3875 |
| *PALB2* (OR≥4) | UKB | 54 | 55 | 66 | 174 | 0 | 0 | 1 | 350 |
|  | CARRIERS | 13 | 10 | 86 | 136 | 0 | 0 | 1 | 246 |
|  | BRIDGES | 26 | 22 | 97 | 262 | 0 | 0 | 1 | 408 |
|  | AMBRY | 83 | 19 | 324 | 470 | 0 | 0 | 1 | 897 |
|  | Combined | 85 | 47 | 429 | 607 | 0 | 0 | 1 | 1169 |
| *ATM*  (OR≥2) | UKB | 136 | 100 | 95 | 467 | 0 | 90 | 1 | 889 |
|  | CARRIERS | 76 | 91 | 109 | 354 | 0 | 0 | 1 | 631 |
|  | BRIDGES | 95 | 123 | 155 | 691 | 0 | 0 | 1 | 1065 |
|  | AMBRY | 285 | 61 | 736 | 1169 | 0 | 0 | 1 | 2252 |
|  | Combined | 364 | 135 | 876 | 1673 | 0 | 14 | 1 | 3063 |
| *CHEK2* (OR≥2) | UKB | 51 | 26 | 18 | 84 | 0 | 22 | 3 | 204 |
|  | CARRIERS | 31 | 32 | 22 | 115 | 0 | 0 | 3 | 203 |
|  | BRIDGES | 38 | 39 | 54 | 150 | 0 | 0 | 2 | 283 |
|  | AMBRY | 111 | 40 | 160 | 270 | 0 | 0 | 2 | 583 |
|  | Combined | 140 | 42 | 194 | 359 | 0 | 6 | 3 | 744 |

#### Supplementary Table 5: All 427 rare missense variants flagged as potentially of reduced penetrance in high penetrance genes *BRCA1*, *BRCA2*, and *PALB2*.

[See separate Supplementary Table 5]

Supplementary Table 6: All 37 rare missense variants flagged as potentially of high penetrance in moderate penetrance genes *ATM* and *CHEK2*. Variants were flagged if they attained a PS4-LLR strength of pathogenic, very strong (P_VSTR) at a target odds of association of OR≥2, and retained a PS4-LLR strength of at least pathogenic, strong (P_STR or P_VSTR) at a target odds of association of OR≥4.

| **Gene** | **HGVS** | **PS4-LLR (Odds≥2)** | **ACMG Evidence Strength (Odds≥2)** | **PS4-LLR (Odds≥4)** | **ACMG Evidence Strength (Odds≥4)** |
| --- | --- | --- | --- | --- | --- |
| ATM | c.6919C>T | 103.9077 | P_VSTR | 47.2335 | P_VSTR |
| ATM | c.7271T>G | 64.52282 | P_VSTR | 57.54294 | P_VSTR |
| ATM | c.1237C>A | 20.68526 | P_VSTR | 20.5814 | P_VSTR |
| ATM | c.8565_8566delinsAA | 20.68526 | P_VSTR | 20.5814 | P_VSTR |
| ATM | c.7875_7876delinsGC | 16.49719 | P_VSTR | 11.55152 | P_VSTR |
| ATM | c.3848T>C | 14.22803 | P_VSTR | 11.65968 | P_VSTR |
| ATM | c.2260C>A | 14.13508 | P_VSTR | 7.315251 | P_STR |
| ATM | c.2446_2447delinsCT | 13.43599 | P_VSTR | 13.20305 | P_VSTR |
| ATM | c.7181C>T | 13.43599 | P_VSTR | 13.20305 | P_VSTR |
| ATM | c.5062A>G | 12.52142 | P_VSTR | 12.26465 | P_VSTR |
| ATM | c.7328G>A | 12.51028 | P_VSTR | 8.437224 | P_VSTR |
| ATM | c.6315G>C | 12.2771 | P_VSTR | 8.076765 | P_VSTR |
| ATM | c.8122G>A | 10.39338 | P_VSTR | 8.480281 | P_VSTR |
| ATM | c.7291A>G | 9.502543 | P_VSTR | 6.011257 | P_STR |
| ATM | c.7355T>C | 9.371261 | P_VSTR | 8.652635 | P_VSTR |
| ATM | c.9022C>T | 9.149022 | P_VSTR | 4.364186 | P_STR |
| ATM | c.2443A>G | 9.12977 | P_VSTR | 7.461982 | P_STR |
| ATM | c.8737G>T | 9.091668 | P_VSTR | 6.858861 | P_STR |
| ATM | c.8194T>G | 8.375619 | P_VSTR | 6.315993 | P_STR |
| ATM | c.3998A>G | 8.29861 | P_VSTR | 8.240711 | P_VSTR |
| ATM | c.2849T>G | 8.210387 | P_VSTR | 4.202597 | P_STR |
| CHEK2 | c.1036C>T | 26.51471 | P_VSTR | 17.62679 | P_VSTR |
| CHEK2 | c.1030A>C | 22.49214 | P_VSTR | 22.4294 | P_VSTR |
| CHEK2 | c.904G>A | 22.47108 | P_VSTR | 20.58012 | P_VSTR |
| CHEK2 | c.751A>T | 21.3448 | P_VSTR | 9.524806 | P_VSTR |
| CHEK2 | c.1270T>C | 21.25351 | P_VSTR | 7.099706 | P_STR |
| CHEK2 | c.1037G>A | 12.44186 | P_VSTR | 7.769077 | P_STR |
| CHEK2 | c.322T>C | 11.77746 | P_VSTR | 9.67211 | P_VSTR |
| CHEK2 | c.350G>A | 11.53711 | P_VSTR | 9.582399 | P_VSTR |
| CHEK2 | c.1180G>A | 10.83872 | P_VSTR | 7.917942 | P_STR |
| CHEK2 | c.1448A>G | 10.34443 | P_VSTR | 7.839179 | P_STR |
| CHEK2 | c.707T>C | 9.308935 | P_VSTR | 7.063863 | P_STR |
| CHEK2 | c.952C>T | 8.956958 | P_VSTR | 4.406319 | P_STR |
| CHEK2 | c.1034A>C | 8.854031 | P_VSTR | 8.517887 | P_VSTR |
| CHEK2 | c.1205_1206delinsTC | 8.474421 | P_VSTR | 7.531757 | P_STR |
| CHEK2 | c.253C>T | 8.298365 | P_VSTR | 8.240459 | P_VSTR |
| CHEK2 | c.608A>G | 8.083902 | P_VSTR | 5.975871 | P_STR |

Supplementary Table 7: Summary of PS4-LLR applied for 26,629 ClinVar missense variants for the five genes of interest. Variants with Likely Benign, Likely Pathogenic, or Pathogenic classifications attained these with at least one star. PS4-LLR calculated using target odds of association for each gene (OR≥4: *BRCA1*, *BRCA2*, *PALB2*; OR≥2: *ATM*, *CHEK2*).

| **Gene** | **ClinVar Classification** | **PS4-LLR towards pathogenicity** | | | | **PS4-LLR not applied/ applicable** | **PS4-LLR towards benignity** | | | | **TOTAL** |
| --- | --- | --- | --- | --- | --- | --- | --- | --- | --- | --- | --- |
|  |  | **vstr** | **str** | **mod** | **sup** |  | **sup** | **mod** | **str** | **vstr** |  |
| ***BRCA1*** | **Pathogenic** | 9 | 6 | 1 | 4 | 40 | 0 | 1 | 1 | 2 | 64 |
|  | **Pathogenic/Likely Pathogenic** | 4 | 1 | 3 | 3 | 37 | 0 | 1 | 2 | 0 | 51 |
|  | **Likely Pathogenic** | 0 | 0 | 0 | 2 | 71 | 0 | 0 | 0 | 0 | 73 |
|  | **Conflicting Classifications** | 8 | 10 | 26 | 22 | 2182 | 18 | 51 | 116 | 198 | 2631 |
|  | **Uncertain Significance** | 2 | 4 | 1 | 7 | 1578 | 4 | 9 | 26 | 43 | 1674 |
|  | **Likely Benign** | 0 | 0 | 0 | 0 | 168 | 0 | 0 | 5 | 4 | 177 |
|  | **Benign/Likely Benign** | 0 | 0 | 0 | 0 | 10 | 0 | 0 | 0 | 5 | 15 |
|  | **Benign** | 0 | 1 | 2 | 2 | 56 | 1 | 6 | 3 | 73 | 144 |
| ***BRCA2*** | **Pathogenic** | 8 | 3 | 1 | 0 | 10 | 0 | 0 | 2 | 0 | 24 |
|  | **Pathogenic/Likely Pathogenic** | 5 | 6 | 1 | 4 | 15 | 1 | 1 | 1 | 0 | 34 |
|  | **Likely Pathogenic** | 1 | 0 | 0 | 1 | 24 | 0 | 0 | 0 | 0 | 26 |
|  | **Conflicting Classifications** | 7 | 28 | 41 | 54 | 3894 | 36 | 109 | 268 | 434 | 4871 |
|  | **Uncertain Significance** | 3 | 13 | 12 | 9 | 3378 | 12 | 34 | 72 | 87 | 3620 |
|  | **Likely Benign** | 0 | 1 | 0 | 0 | 206 | 1 | 1 | 5 | 4 | 218 |
|  | **Benign/Likely Benign** | 0 | 0 | 0 | 1 | 15 | 0 | 2 | 0 | 6 | 24 |
|  | **Benign** | 1 | 1 | 0 | 1 | 66 | 0 | 2 | 3 | 61 | 135 |
| ***PALB2*** | **Pathogenic** | 0 | 0 | 0 | 0 | 0 | 0 | 1 | 0 | 0 | 1 |
|  | **Pathogenic/Likely Pathogenic** | 0 | 0 | 0 | 0 | 0 | 0 | 0 | 0 | 0 | 0 |
|  | **Likely Pathogenic** | 0 | 0 | 0 | 0 | 2 | 0 | 0 | 0 | 0 | 2 |
|  | **Conflicting Classifications** | 2 | 1 | 17 | 8 | 252 | 10 | 22 | 45 | 110 | 467 |
|  | **Uncertain Significance** | 2 | 6 | 14 | 28 | 2162 | 5 | 63 | 65 | 102 | 2447 |
|  | **Likely Benign** | 0 | 0 | 1 | 0 | 30 | 0 | 0 | 0 | 0 | 31 |
|  | **Benign/Likely Benign** | 0 | 0 | 0 | 0 | 6 | 0 | 0 | 0 | 0 | 6 |
|  | **Benign** | 0 | 0 | 0 | 0 | 5 | 1 | 0 | 0 | 0 | 6 |
| ***ATM*** | **Pathogenic** | 1 | 0 | 0 | 0 | 5 | 0 | 1 | 0 | 1 | 8 |
|  | **Pathogenic/Likely Pathogenic** | 5 | 2 | 3 | 1 | 15 | 0 | 1 | 3 | 0 | 30 |
|  | **Likely Pathogenic** | 3 | 1 | 2 | 1 | 29 | 1 | 2 | 0 | 0 | 39 |
|  | **Conflicting Classifications** | 7 | 17 | 32 | 30 | 386 | 21 | 48 | 71 | 118 | 730 |
|  | **Uncertain Significance** | 7 | 39 | 83 | 121 | 6322 | 65 | 197 | 203 | 117 | 7154 |
|  | **Likely Benign** | 0 | 0 | 1 | 1 | 53 | 0 | 1 | 0 | 1 | 57 |
|  | **Benign/Likely Benign** | 0 | 0 | 0 | 0 | 14 | 0 | 0 | 0 | 2 | 16 |
|  | **Benign** | 0 | 0 | 0 | 0 | 13 | 0 | 0 | 0 | 2 | 15 |
| ***CHEK2*** | **Pathogenic** | 0 | 0 | 0 | 0 | 0 | 0 | 0 | 0 | 0 | 0 |
|  | **Pathogenic/Likely Pathogenic** | 1 | 0 | 0 | 0 | 0 | 0 | 0 | 0 | 2 | 3 |
|  | **Likely Pathogenic** | 1 | 0 | 0 | 1 | 3 | 0 | 0 | 0 | 0 | 5 |
|  | **Conflicting Classifications** | 12 | 4 | 5 | 7 | 42 | 5 | 10 | 10 | 23 | 118 |
|  | **Uncertain Significance** | 11 | 22 | 21 | 51 | 1459 | 7 | 52 | 52 | 28 | 1703 |
|  | **Likely Benign** | 0 | 0 | 0 | 0 | 8 | 0 | 0 | 0 | 0 | 8 |
|  | **Benign/Likely Benign** | 0 | 0 | 0 | 0 | 2 | 0 | 0 | 0 | 0 | 2 |
|  | **Benign** | 0 | 0 | 0 | 0 | 0 | 0 | 0 | 0 | 0 | 0 |

Supplementary Table 8: 2,030 *BRCA1*/*BRCA2* missense variants (with VUS or conflicting classifications in ClinVar) annotated with PS4-LLR, functional data, and in silico predictive tool scores. Each variant has been assigned an overall classification using a) just functional and predictive data, and b) using case-control, functional, and predictive data. At least two pieces of concordant evidence were required for a classification, otherwise the variant was assigned a default classification of VUS. A target odds of association of OR≥4 was used to calculate PS4-LLR for *BRCA1* and *BRCA2*.

[See separate Supplementary Table 8]

Supplementary Table 9: Meta-analysed OR for 926 rare missense variants present in at least two of the three unselected datasets (UK Biobank, BRIDGES, CARRIERS). Where variant carriers were identified in cases and not controls (cases≥1, controls=0), the Haldane-Anscombe correction was applied (+0.5 to all cells). Where variant carriers were identified in controls and not cases (cases=0, controls≥1), data was not included in final meta-analysis.

[See separate Supplementary Table 9]

Supplementary Table 10: Exemplar threshold differences between total cases and total controls (before false positive/negative results) at a range of target odds of association, calibrated at 1, 2, or 3 observations in either cases or controls.

| **Target odds of association** | **Scenario** | | **Threshold difference between total cases and controls** |
| --- | --- | --- | --- |
|  | **No. case carriers** | **No. control carriers** |  |
| OR>2 | 0 | 1 | **3.45x number of controls** |
| OR>2 | 0 | 2 | **5.50x number of controls** |
| OR>2 | 0 | 3 | **7.55x number of controls** |
| OR>2 | 1 | 0 | **1.7x number of cases** |
| OR>2 | 2 | 0 | **2.75x number of cases** |
| OR>2 | 3 | 0 | **3.8x number of cases** |
| OR>3 | 0 | 1 | **4.25x number of controls** |
| OR>3 | 0 | 2 | **6.85x number of controls** |
| OR>3 | 0 | 3 | **9.45x number of controls** |
| OR>3 | 1 | 0 | **1.4x number of cases** |
| OR>3 | 2 | 0 | **2.30x number of cases** |
| OR>3 | 3 | 0 | **3.15x number of cases** |
| OR>4 | 0 | 1 | **4.95x number of controls** |
| OR>4 | 0 | 2 | **8.00x number of controls** |
| OR>4 | 0 | 3 | **11.1x number of controls** |
| OR>4 | 1 | 0 | **1.25x number of cases** |
| OR>4 | 2 | 0 | **2.00x number of cases** |
| OR>4 | 3 | 0 | **2.75x number of cases** |
| OR>6 | 0 | 1 | **6.2x number of controls** |
| OR>6 | 0 | 2 | **10.10x number of controls** |
| OR>6 | 0 | 3 | **14.05x number of controls** |
| OR>6 | 1 | 0 | **1.05x number of cases** |
| OR>6 | 2 | 0 | **1.70x number of cases** |
| OR>6 | 3 | 0 | **2.35x number of cases** |
| OR>8 | 0 | 1 | **7.3x number of controls** |
| OR>8 | 0 | 2 | **11.95x number of controls** |
| OR>8 | 0 | 3 | **16.65x number of controls** |
| OR>8 | 1 | 0 | **0.9x number of cases** |
| OR>8 | 2 | 0 | **1.50x number of cases** |
| OR>8 | 3 | 0 | **2.1x number of cases** |
| OR>10 | 0 | 1 | **8.25x number of controls** |
| OR>10 | 0 | 2 | **13.60x number of controls** |
| OR>10 | 0 | 3 | **19.05x number of controls** |
| OR>10 | 1 | 0 | **0.85x number of cases** |
| OR>10 | 2 | 0 | **1.35x number of cases** |
| OR>10 | 3 | 0 | **1.9x number of cases** |
| OR>100 | 0 | 1 | **32.20x number of controls** |
| OR>100 | 0 | 2 | **58.20x number of controls** |
| OR>100 | 0 | 3 | **85.70x number of controls** |
| OR>100 | 1 | 0 | **0.3x number of cases** |
| OR>100 | 2 | 0 | **0.60x number of cases** |
| OR>100 | 3 | 0 | **0.85x number of cases** |
| OR>1000 | 0 | 1 | **136.70x number of controls** |
| OR>1000 | 0 | 2 | **281.35x number of controls** |
| OR>1000 | 0 | 3 | **444.20x number of controls** |
| OR>1000 | 1 | 0 | **0.15x number of cases** |
| OR>1000 | 2 | 0 | **0.30x number of cases** |
| OR>1000 | 3 | 0 | **0.45x number of cases** |

Supplementary Table 11: Summary of 4,690 rare missense variants attaining PS4-LLR evidence at different target odds of association, calculated for OR≥2 and OR≥4 for all genes

|  |  | **Target odds of association** | |
| --- | --- | --- | --- |
| **Gene** |  | **OR≥2** | **OR≥4** |
| ***BRCA1*** | **P_VSTR** | 32 | 25 |
|  | **P_STR** | 44 | 24 |
|  | **P_MOD** | 71 | 35 |
|  | **P_SUP** | 82 | 43 |
|  | **NA** | 86 | 102 |
|  | **B_SUP** | 45 | 22 |
|  | **B_MOD** | 116 | 72 |
|  | **B_STR** | 171 | 159 |
|  | **B_VSTR** | 161 | 326 |
| ***BRCA2*** | **P_VSTR** | 42 | 26 |
|  | **P_STR** | 83 | 54 |
|  | **P_MOD** | 140 | 59 |
|  | **P_SUP** | 148 | 72 |
|  | **NA** | 167 | 195 |
|  | **B_SUP** | 104 | 52 |
|  | **B_MOD** | 261 | 155 |
|  | **B_STR** | 323 | 362 |
|  | **B_VSTR** | 302 | 595 |
| ***PALB2*** | **P_VSTR** | 6 | 4 |
|  | **P_STR** | 21 | 8 |
|  | **P_MOD** | 51 | 35 |
|  | **P_SUP** | 51 | 38 |
|  | **NA** | 84 | 47 |
|  | **B_SUP** | 30 | 16 |
|  | **B_MOD** | 94 | 88 |
|  | **B_STR** | 120 | 112 |
|  | **B_VSTR** | 104 | 213 |
| ***ATM*** | **P_VSTR** | 24 | 14 |
|  | **P_STR** | 61 | 34 |
|  | **P_MOD** | 125 | 74 |
|  | **P_SUP** | 154 | 47 |
|  | **NA** | 135 | 191 |
|  | **B_SUP** | 87 | 50 |
|  | **B_MOD** | 254 | 221 |
|  | **B_STR** | 290 | 234 |
|  | **B_VSTR** | 245 | 510 |
| ***CHEK2*** | **P_VSTR** | 25 | 8 |
|  | **P_STR** | 27 | 18 |
|  | **P_MOD** | 28 | 18 |
|  | **P_SUP** | 60 | 44 |
|  | **NA** | 42 | 46 |
|  | **B_SUP** | 12 | 6 |
|  | **B_MOD** | 63 | 50 |
|  | **B_STR** | 66 | 53 |
|  | **B_VSTR** | 53 | 133 |

Supplementary Table 12: Alternative target odds of association for each gene, where all 13,966 variants are presented with PS4-LLR calculated for a range of odds (OR≥2, OR≥2, and a bespoke OR for each gene calculated in a previous large meta-analysis, where OR≥8.73 for *BRCA1*, OR≥5.68 for *BRCA2*, OR≥4.30 for *PALB2*, OR≥2.17 for *ATM*, and OR≥2.44 for *CHEK2*).

[See separate Supplementary Table 12]

Supplementary Table 13: List of *BRCA1*, *BRCA2*, and *PALB2* PTVs with Pathogenic or Likely Pathogenic ClinVar classifications and strong or very strong Benign PS4-LLR evidence at OR≥4. The PS4-LLR evidence at OR≥2 is also provided to demonstrate weakening of benign evidence at lower target odds of association.

[See separate Supplementary Table 13]

#### Supplementary Table 14: Summary of rules defined for inclusion of data from case-control datasets in case-control analysis using the PS4-LR-Calculator.

| **Rule** | **Reason implemented** | **Application of rule** |
| --- | --- | --- |
| Quantify enrichment of pathogenic variants in enriched case series | Clinically collected national laboratory data is likely to be enriched for pathogenic variant carriers due to restrictive eligibility criteria for testing compared to studies which did not select for ascertainment. This means findings are not directly comparable. | Enrichment of pathogenic variants can be estimated using the relative frequency of putatively pathogenic rare protein truncating variants (PTVs) in each dataset, defined as variants which met the criteria for PVS1 and did not meet the criteria for BA1 per the guidelines and frequency thresholds defined by the respective gene variant curation expert panel.  For a given gene ($g$), the rare PTV allele frequency ($AF$) was calculated for each unselected case series. These frequencies were then averaged to give a per-gene AF for unselected breast cancer datasets (${gAF}_{unselected}$). The allele frequency was similarly calculated for each laboratory case dataset (${gAF}_{enriched}$).  The gene-specific enrichment factor ($gEF$) was thus calculated for each enriched dataset (NDRS, Ambry) (Supplementary Table 1), essentially calculating the risk ratio for rare PTVs in the enriched dataset:  $gEF=\frac{{gAF}_{enriched}}{{gAF}_{unselected}}$  Lower ($LCI$) and upper ($UCI$) 95% confidence intervals were also calculated for each value of $gEF$:  $LCI=e^{\log\left( gEF \right)-\left( 1.96*\surd\left( \frac{1}{a}-\frac{1}{c}+\frac{1}{b}-\frac{1}{d} \right) \right)}$; $UCI=e^{\log\left( gEF \right)+\left( 1.96*\surd\left( \frac{1}{a}-\frac{1}{c}+\frac{1}{b}-\frac{1}{d} \right) \right)}$  a = number of PTV carriers in gene $g$ in enriched dataset  c = total persons tested for gene $g$ in enriched dataset  b = number of PTV carriers in gene $g$ in all three unselected datasets  d = total persons tested for gene $g$ in all three unselected datasets  To incorporate this enrichment factor into the PS4-LR-Calc, the lower boundary for the hypothesis of association ($p_{l}$) was adjusted by the EF as follows:  $p_{l}= \frac{(gEFx)c}{(gEFx)c+d}$  where the threshold (target) odds ratio ($x$) was multiplied by the EF. For *BRCA1*, *BRCA2*, and *PALB2*, $x$=4. For *ATM* and *CHEK2*, $x$=2. When likelihood was calculated for the unselected datasets, $gEF$=1, therefore having no impact.  In this way, the minimum threshold for the hypothesis of association is raised for enriched datasets by the factor we estimate the dataset to be enriched, which ultimately means more case observations are needed to meet this threshold and assign pathogenic evidence. This accounted for the measured increase in pathogenic variant observations compared to unselected datasets, and allowed for direct combination of the final likelihood ratio for each dataset. |
| Remove single observations only seen in one dataset | Single observations not repeated in multiple datasets are vulnerable to potential technical error from erroneous variant calling, and risk over-estimation of evidence strength resulting from the underlying binomial distribution when n=1. | LR produced from a single observation in a single dataset is disregarded. LR produced from single observations in multiple datasets are combined as normal. This approach is consistent with the direction of ClinGen groups such as the low-risk allele working group, and with other groups researching quantitation of case-control evidence such as the Zanti *et al*. approach, which require at least 3 observations of a variant before quantitative approaches are used for case-control data.^28,29^ |
| Laboratory data must have equivalent, matched controls | A control set of observations is essential to perform a case:control calculation. Ethnicity-matching (and if possible location-matching) is also essential due to differing frequencies in variant frequency in different ancestral populations, particularly for founder variants. | For the NDRS case dataset, the control cohort from UKB (419,373 controls) was proportionally stochastically split between the 18,477 UKB cases and 44,917 NDRS cases. Maximising the number of controls in each partition while minimising the case:control ratio, 122,232 controls were retained for UKB cases and 297,141 used with NDRS cases (a ratio of 1:6.62 cases:controls for each) (Supplementary Figure 4)  All variant carriers in the UKB control series were randomly sampled between the NDRS and UKB datasets according to the same proportional split (29.1% probability to be in UKB control cohort, 70.8% probability to be in UK laboratory control cohort). NDRS cases were merged with the proportional split of UKB controls using gene and coding DNA nomenclature.  We acknowledge that stochastic splitting of the UKB control series in this way potentially compounds any UKB-specific effects across two sets of case series (from NDRS and from UKB). However, at this time there are no other large, ethnicity- and location-matched control series available to use with the NDRS case series.  For the Ambry case dataset, Ambry cases were merged with gnomAD v4.1.0 exome data using the gene and coding DNA nomenclature provided post-liftover to build 37. Notably, gnomAD data are population controls, not non-breast controls. |
| Evidence towards benignity may not be applied where the dataset is known to be depleted for benign variants | Benign and likely benign variants data is not routinely submitted from English laboratories to NDRS. Variant counts for these variants are therefore likely depleted, and evidence towards benignity would be inflated as a result. | After assigning equivalent matched controls from UK Biobank, evidence towards benignity (PS4-LLR < 0) in the NDRS dataset was excluded from dataset combination. Variants which were only present in the NDRS/UK Biobank dataset and which attained evidence towards benignity were excluded from analysis. |
| Constrain proportional size of cases and controls | Imbalance of cases and controls tested in the datasets led to application of pathogenic evidence when carriers were only identified in controls (i.e. false positive results), or benign evidence when carriers were only identified in cases (i.e. false negative results). This occurred when carrier count was low (<3). | False positive and false negative results only began to appear in the dataset when the size of the difference between total cases and total controls in the case:control dataset breached a certain threshold. This threshold difference is specific to a) the number of carriers, and b) the target odds of association. Therefore, we could calculate the threshold difference for a range of example target odds of association when there were 1, 2, or 3 carriers (Supplementary Table 10).  Threshold difference was calculated using a synthetic dataset comprising 5000 scenarios of different case:control sizes (where the smallest scenario was a dataset of 500 controls and 10,000 cases, and the largest was a dataset of 500,000 controls and 10,000 cases). For each scenario, the ratio between controls:cases was calculated (again, the smallest being 0.05:1 and the largest being 50:1). Each scenario was then assigned 1, 2, or 3 carriers in either cases or controls (with the other value set to 0, i.e. where 1 control carrier was assigned, 0 case carriers were assigned).  For each scenario, PS4-LLR was then calculated. The scenario where PS4-LLR was closest to 0 was set as the maximum threshold difference before false positive/negative results are calculated (e.g. PS4-LLR being >0 for a scenario where variants were only seen in controls) (Supplementary Figure 5). Interestingly, we found that at higher target odds of association, a larger number of controls compared to cases were necessary to avoid false negative outputs for variants only seen in cases (Supplementary Table 10).  For the actual datasets, all possible threshold differences were calculated for at every target odds of association when there were 1,2, or 3 carriers in either cases or controls. Where the number of carriers and the target odds of association caused the difference between number of cases and controls to exceed the threshold difference (thus producing a false positive/negative result), the data for that variant in that dataset was not used in the combined LR calculation. |
| Restrict inclusion by observed case-control signal | Variants with an observed odds ratio between the target odds of association and non-association (and with 95% confidence intervals also within these limits) have a <5% chance of the true underlying likelihood being more extreme than either target odds, and result in the total likelihood space (the percentage of the area under the binomial distribution being covered by each target odds) being very small. In these scenarios, slight differences in the number of case and control carriers can translate to inappropriately large likelihood estimates.  It is highly likely these variants are of reduced penetrance compared to the standard target odds of association. | To explore this concept, the OR, LR, and likelihood space was calculated for every possible combination of 0-100 cases and 0-100 controls using a target odds of association of 4 and testing denominators based on the BRIDGES dataset (Supplementary Figure 6a). Variants with an observed OR and 95% confidence intervals between the target odds of non-association (1) and target odds of association (4) form a group with the smallest likelihood space (Supplementary Figure 6b).  18 missense and 2 protein truncating variants in the combined dataset met this definition and were therefore excluded from downstream analysis:   - 4 *BRCA1* variants (c.4910C>T p.(Pro1637Leu), c.314A>G p.(Tyr105Cys), c.2315T>C p.(Val772Ala), c.2596C>T p.(Arg866Cys)) - 10 *BRCA2* variants (c.3545_3546del p.(Phe1182Ter), c.3326C>T p.(Ala1109Val), c.223G>C p.(Ala75Pro), c.3568C>T p.(Arg1190Trp), c.8917C>T p.(Arg2973Cys), c.6853A>G p.(Ile2285Val), c.6317T>C p.(Leu2106Pro), c.1964C>G p.(Pro655Arg), c.3515C>T p.(Ser1172Leu), c.1889C>T p.(Thr630Ile)) - 1 *PALB2* variant (c.656A>G p.(Asp219Gly)) - 1 *ATM* variant (c.4388T>G p.(Phe1463Cys)) - 4 *CHEK2* variants (c.444+1G>A, c.1312G>T p.(Asp438Tyr), c.1427C>T p.(Thr476Met), and previously reported low-risk allele *CHEK2* c.470T>C p.(Ile157Thr)^13^) |

Supplementary Table 15: All functional study data used in application of PS3 and BS3, evidence application for each variant as recommended by the ClinGen ENIGMA *BRCA1* and *BRCA2* Variant Curation Expert Panel (VCEP).

| **Study** | **Gene** | **PMID** | **Assay output (Functional)** | **Assay output (Loss of Function)** |
| --- | --- | --- | --- | --- |
| Bouwman et al., 2020 | *BRCA1* | 32546644 | 3x neutral, or 2x neutral and 1x intermediate | 3x deleterious, or 2x deleterious + 1x intermediate result |
| Findlay et al., 2018 | *BRCA1* | 30209399 | >-0.748 | <-1.328 |
| Fernandes et al., 2019 | *BRCA1* | 30765603 | Mean ≥80% wild type activity | Mean <80% wild type activity |
| Petitalot et al., 2019 | *BRCA1* | 30257991 | 1P | 3P |
| Starita et al., 2018 | *BRCA1* | 30219179 | Zero depleted replicates | 3-4 depleted replicates |
| Bouwman et al., 2013 | *BRCA1* | 23867111 | Neutral | Deleterious |
| Mesman et al., 2019 | *BRCA2* | 29988080 | Complementation shown | Complementation not shown |
| Richardson et al., 2021 | *BRCA2* | 33609447 | >2.25 | <1.78 |
| Ikegami et al., 2020 | *BRCA2* | 32444794 | Class 1 or 2 | Class 4 or 5 |
| Biswas et al., 2020 | *BRCA2* | 33293522 | P ≤ 0.05 | P > 0.99 |
| Hart et al., 2019 | *BRCA2* | 29884841 | FC ≥ 2.41 | FC ≤ 1.66 |
